# Health journal coverage of climate change and health: a bibliometric study

**DOI:** 10.1101/2023.10.19.23297267

**Authors:** Joy Muhia, Melissa L. Rethlefsen, Ben Rossington, Florence Wedmore, Anandita Pattnaik, Richard Smith, Sara Schroter

## Abstract

**Objectives:** To find what proportion of a broad set of health journals have published on climate change and health, how many articles they have published, and when they first published on the subject.

**Design:** Bibliometric study.

**Setting and particbipants:** We conducted electronic searches in Ovid MEDLINE ALL for articles about climate change and human health published from 1860 to 31 December 2022 in 330 health journals. There were no limits by language or publication type. Results were independently screened by two raters for article eligibility.

**Results:** After screening there were 2932 eligible articles published across 253 of the 330 journals between 1947 and 2022; most (2795/2932; 95%) were published in English. A few journals published articles in the early 90s, but there has been a rapid increase since about 2006. We were unable to categorise the types of publication but estimate that fewer than half are research papers. While articles were published in journals in 39 countries, two-thirds (1929/2932; 66%) were published in a journal published in the UK or the US. Almost a quarter (77/330; 23%) of the journals published no eligible articles, and almost three-quarters (241/330; 73%) published five articles or fewer. The publication of joint editorials in over 200 journals in 2021 and 2022 boosted the number of journals publishing something on climate change and health. A third of the (112/330; 34%) journals in our sample published at least one of the joint editorials, and almost a third of those (32/112; 29%) were publishing on climate change and health for the first time.

**Conclusions:** Health journals are rapidly increasing the amount they publish on climate change and health, but despite climate change being the major threat to global health many journals had until recently published little or nothing. A joint editorial published in multiple journals increased coverage, and for many journals it was the first thing they published on climate change and health.

**Strengths and limitations of this study:**

1. We looked at coverage of climate change at both the article and journal level from 1860 to 2022; previous bibliometric studies have analysed results only at the article level and in restricted time periods
2. Two independent raters screened the title and abstract (where available) of every article identified by the MEDLINE search to assess eligibility
3. Journals not currently indexed in MEDLINE had to be excluded as hand searching was not feasible for such a large sample of journals
4. It’s important to be cautious in generalising to all health journals from our sample
5. Our study included analysis of an intervention (publication of the joint editorials in multiple journals) to increase coverage on climate change and health in health journals.

## INTRODUCTION

“Climate change is the biggest global health threat of the 21st century,” concluded the Lancet Commission on Climate Change in 2009.[1] Harm arises from heatwaves, storms, fires, floods, shortages of water, food, and shelter, changing patterns of disease, forced migration, and conflict.[1] Yet if the world were to make the changes necessary to respond to climate change—for example, ending the use of fossil fuels, shifting diets to being largely plant-based, and encouraging active transport, then the change could also be “the greatest opportunity for global health.”[2]

But a series of Lancet Countdowns on Climate Change, the latest in 2022, show that the global temperature and emissions continue to rise, and the global response is inadequate.[3] Health systems, like all other human activities, emit greenhouse gases, and if health systems were a country they would be the fifth biggest emitter.[4] Most health systems have rising emissions of greenhouse gases.[3] The NHS in England is responsible for between 4 and 5% of UK global emissions and is committed to achieving carbon net zero by 2040 on all that it controls directly and by all that it consumes by 2045.[5] All other health systems will also have to achieve net zero by 2050 if the world is to respond adequately to climate change, and the changes will have to be radical, affecting all parts of the system including all medical specialties. Indeed, nobody knows exactly how health systems can achieve net zero, and much research will be needed. In addition, health systems must adapt to the climate changes that have already occurred and will continue to worsen for a while even if global emissions are cut.

The seriousness of the threat to health and the necessity for health systems and medical practice to change mean that climate change should receive extensive coverage in health journals.

Climate change from rising carbon dioxide levels was predicted as long ago as the late 19^th^ century. In 1988, the head of the NASA Goddard Institute for Space Studies told a US Senate Committee: “The greenhouse effect has been detected, and it is changing our climate now.”[6] The first Earth Summit was held in Rio de Janeiro in 1992.

Health journals could have been covering climate change and health for many decades. The Lancet Countdown on Climate Change conducted a bibliometric study of climate change and health and showed a near exponential growth in the number of peer reviewed articles between 1985 and 2022, with fewer than 100 in 2000 and over 3200 in 2021.[3] There was a 22% increase between 2020 and 2021.[3] A supporting study of reports between 2013 and 2020 showed that more than four fifths of the articles (86%) covered the implications for health of climate change rather than mitigation or adaptation.[7] The authors used supervised machine learning and other natural language processing methods, did not review individual studies, and did not conduct an analysis by journal.[7] Authors of a study published in 2016 searched in PubMed and Science Direct for publications from 1990 to 2014 for articles that contained one climate term and one health term in either the title, abstract, or as a Medical Subject Heading (MeSH) term.[8] The authors found few articles until 2006 and then an exponential increase, but papers dealing with health and climate were about half that of climate related articles in other sectors.

Another group searched the National Institute of Environmental Health Sciences Climate Change and Human Health Literature Portal, which includes global peer-reviewed research and grey literature conducted in North America (USA, Mexico, and Canada).[9][10] The database was compiled by searching PubMed, Web of Science, and Google Scholar for research on climate change and health published from 2012 to 2019. The group reviewed individual articles and identified 7082 original research articles published between 2012 and 2019 with an average annual increase of 23%.

These previous bibliometric studies dealt with articles not journals, and only one study reviewed individual articles to ensure that they did cover climate change and health—and that study covered only the years after 2012 and research conducted in North America.[9, 10] Terminology for this topic has changed over the years and simply mentioning climate change doesn’t mean that the article is addressing the topic. We set out to find the extent of the coverage of climate change in health journals over time in a broad set of health journals, using a broad search strategy with two raters screening the eligibility of the search results. We were particularly interested to know when and which health journals first published on climate change and health and how many had not published anything on the subject.

We were prompted to do this by a programme developed by the UK Health Alliance on Climate Change to encourage over 200 health journals around the world to publish the same editorials on climate change and health before COP26 and COP27, the United Nations annual meetings on climate change, in 2021 and 2022.[11, 12] The aim of the editorials was to encourage more action by governments and to reach more health professionals with information on the importance for health of acting on climate change. Before conducting our study, we had the impression that many of the journals had published little or nothing on climate change until they published these editorials.

## METHODS

### Data sources and searches

We conducted electronic searches in Ovid MEDLINE ALL for articles about climate change published from 1860 to 31 December 2022.

### Search: Identification of journals

To identify a broad range of health journals to include in the search and to avoid retrieving articles in journals that focus on environmental health rather than climate change, we took three purposive journal samples and pooled the results. We identified the top 50 most highly cited clinical medicine journals based on impact factor from Clarivate’s Journal Citation Reports Database (JCR 2021); all journals listed in the JCR 2021 category of “Medicine, General & Internal”; and all journals that published the joint editorial coordinated by UKHACC, titled “*Call for emergency action to limit global temperature increases, restore biodiversity, and protect health”* in 2021. After pooling the three samples, we removed duplicated journals and excluded journals that were not indexed in MEDLINE in January 2023.

To ensure a comprehensive list of journals meeting inclusion criteria, each journal was individually reviewed in the NLM Journals Database (https://www.ncbi.nlm.nih.gov/nlmcatalog/journals/). Each included journal was reviewed for title changes since being indexed in MEDLINE. When title changes were identified, each previous and current journal title was reviewed to see whether it was indexed in MEDLINE. Published journals that were not labelled “Currently indexed by MEDLINE” were excluded. For currently published titles indexed in MEDLINE, all journal titles for the same publication were included in the search to ensure maximum coverage of each journal if they were or had been indexed in MEDLINE. To be considered the same publication, volume numbering needed to continue across the title changes (e.g. British Medical Journal (Clinical Research Edition) ceased with volume 296, number 6639 and BMJ (Clinical Research Edition) began with volume 297, number 6640).

### Search: Identification of articles on climate change and human health

Medical Subject Headings (MeSH) have not been available for the topic of climate change until 2010 and an array of terms have been used to describe the topic. We created a list of keywords based on those used in previous studies[13–18] and supplemented this with additional keywords we thought should also be included following discussions as a team and experimenting with different keywords. We tested the initial search strategy on a single journal (*The BMJ*) and the research team reviewed the results to help decide whether the search should be more or less specific.

We found the inclusion of keywords for extreme weather conditions led to the inclusion of some ineligible articles but their exclusion from the search would lead to the exclusion of some relevant articles. Based on the pilot results and to estimate the magnitude of the problem in the full set of articles from all journals we decided to conduct two separate literature searches: 1) the “main search” with the extreme weather-related terms excluded (see Appendix 1 Line 409) and 2) the “extreme weather conditions search” with additional weather-related terms that were not included in the main search (see Appendix 1 Line 410).

### Data extraction and synthesis

The main literature search was conducted in Ovid MEDLINE All on 26 January 2023 and updated on 31 March 2023. Results were exported to Covidence systematic review software (Veritas Health Innovation, Melbourne, Australia. Available at www.covidence.org) and Microsoft Excel. For every article identified by the main search, two independent raters screened articles for eligibility (using the inclusion and exclusion criteria in Appendix 2) based on article title and abstract only. For four non-English articles where no title or abstract were included from the export from MEDLINE, raters used the full text article link available in Covidence to access the full text of the article to assess the article title and or abstract for eligibility. All conflicting ratings were resolved independently by a third rater on Covidence. Ineligible articles were then removed from the Excel file which was exported from the search results in MEDLINE. For the “extreme weather conditions” search, the search was conducted on 28 January 2023. We took a random sample of 5% articles from the search results and screened them for eligibility in the same way as for the main search.

### Exclusion and inclusion criteria

For the search, all articles in the included journals available in MEDLINE and published 1860-2022 were eligible; there were no limits by language or publication type. Appendix 2 gives details of the inclusion/exclusion criteria applied by raters.

### Statistical analysis

Descriptive statistics were used to describe the article language, year of publication, journal name, and journal country of publication.

### Patient and public involvement

BR, who completed a placement at the UK Health Alliance on Climate Change in autumn 2022 as part of the NHS Graduate Management Training Scheme, is our public representative and has been involved from the start of the study and at every stage of the study and in the writing of the manuscript.

## RESULTS

### Sample

We included a sample of 330 journals from across the three sources (Figure 1). The main search strategy identified a total of 3571 articles published. Five duplicates were removed after identification in Covidence and two during data analysis. After screening by raters 2932 (82%) of the remaining 3564 articles were considered eligible for inclusion.

**Figure 1:**
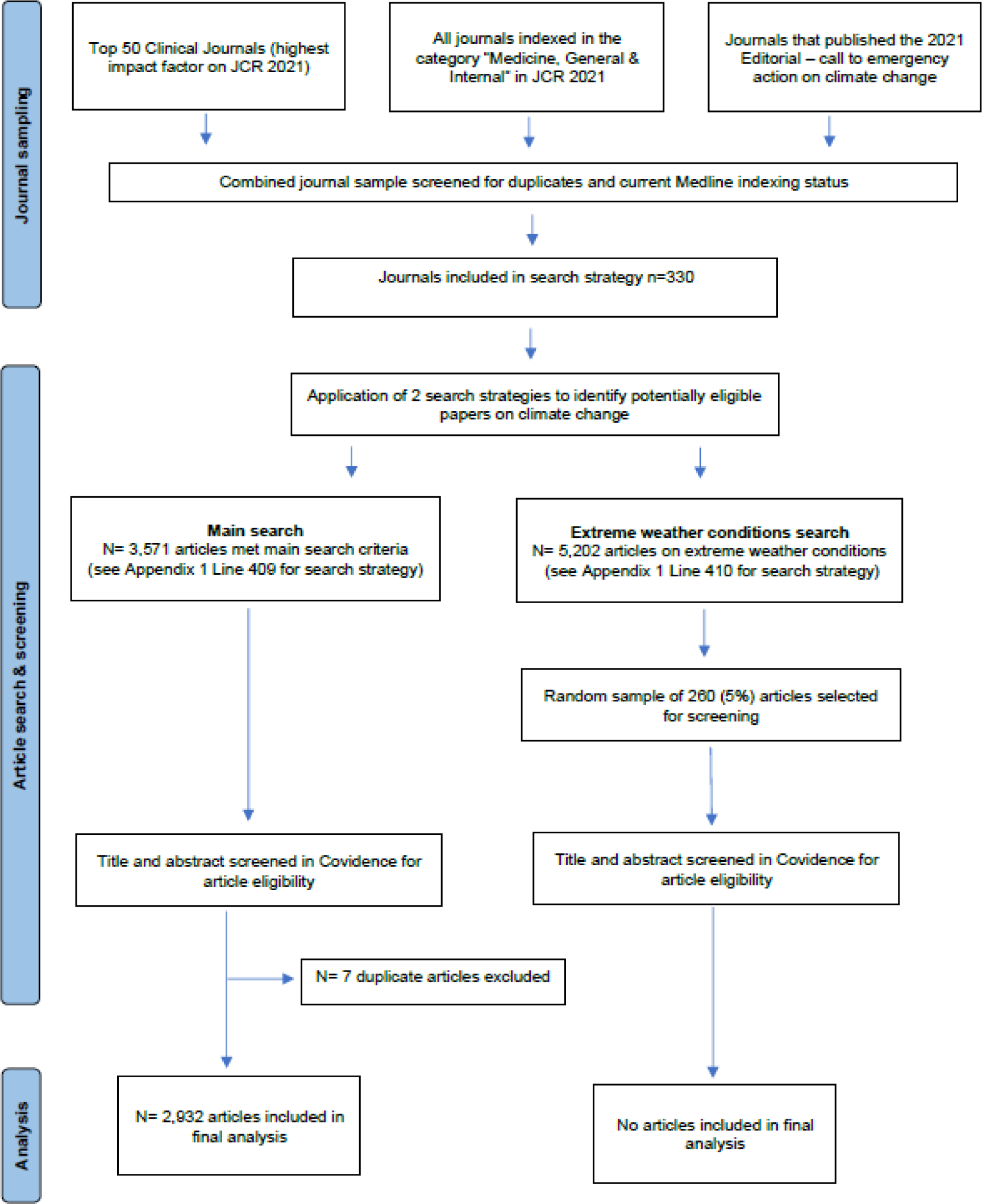
Flow chart of included articles.

The extreme weather conditions search identified 5202 articles. Screening of a random sample of 260 (5%) of these identified that only 6 (2%) were eligible for inclusion so we decided not to screen the results from the entire extreme weather conditions search and not to include any articles from this search in the final data analysis.

### Distribution over time

Figure 2 shows the number of articles on climate change and health published each year between 1947, the earliest year of publication we identified, and 2022. We did not identify any articles published from before 1947. Figure 3 shows the cumulative frequency of articles published between 1947 and 2022. There were occasional articles published in the 1990s, but the increase in the number of articles began in the early 2000s. Fewer than 100 articles had been published before 2000, slightly more than 600 by 2010, around 2000 by 2020, and almost 3000 by 2022.

**Figure 2:**
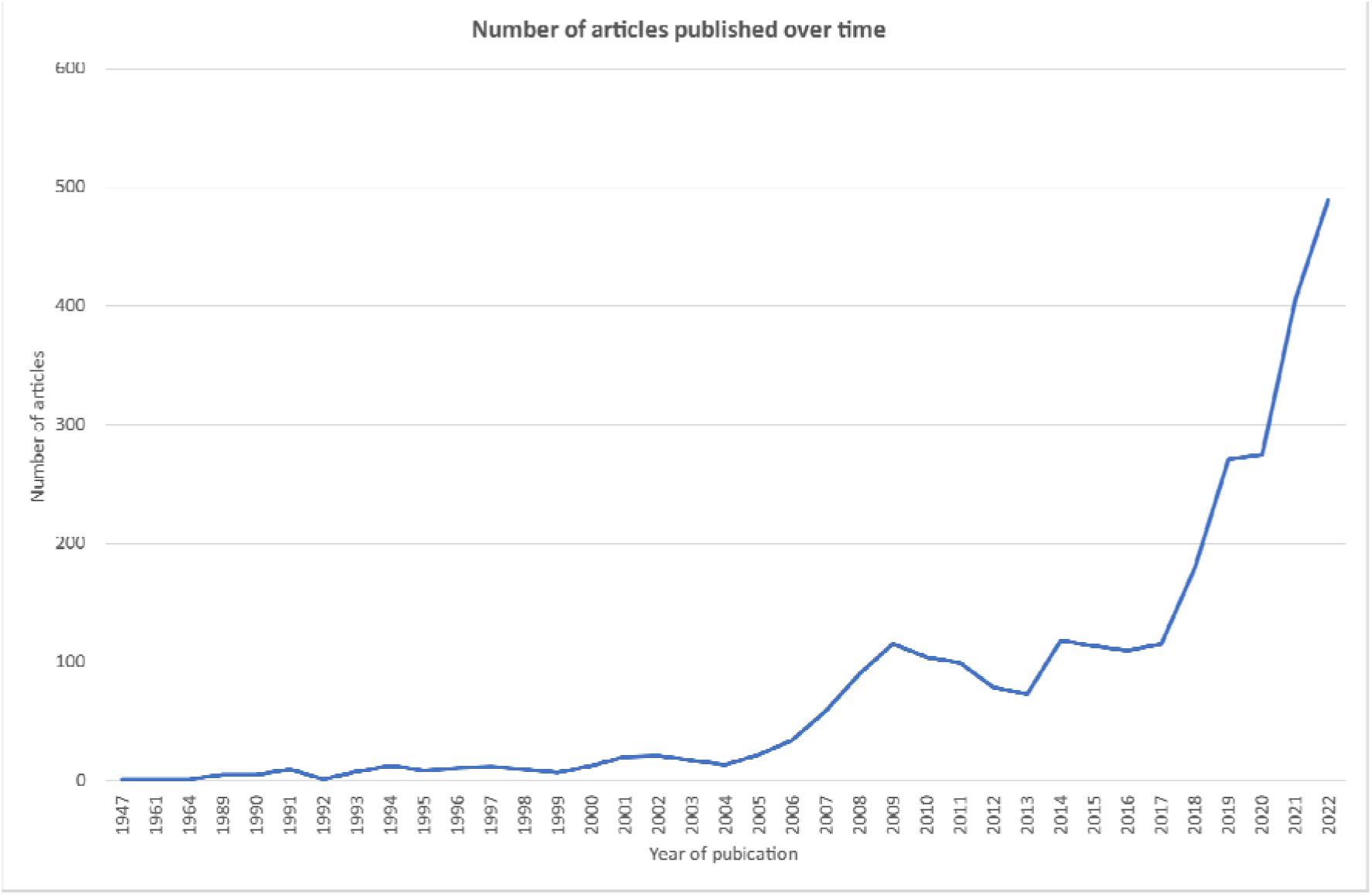
Number of articles published each year between 1947 and 2022 (n=2932)

**Figure 3:**
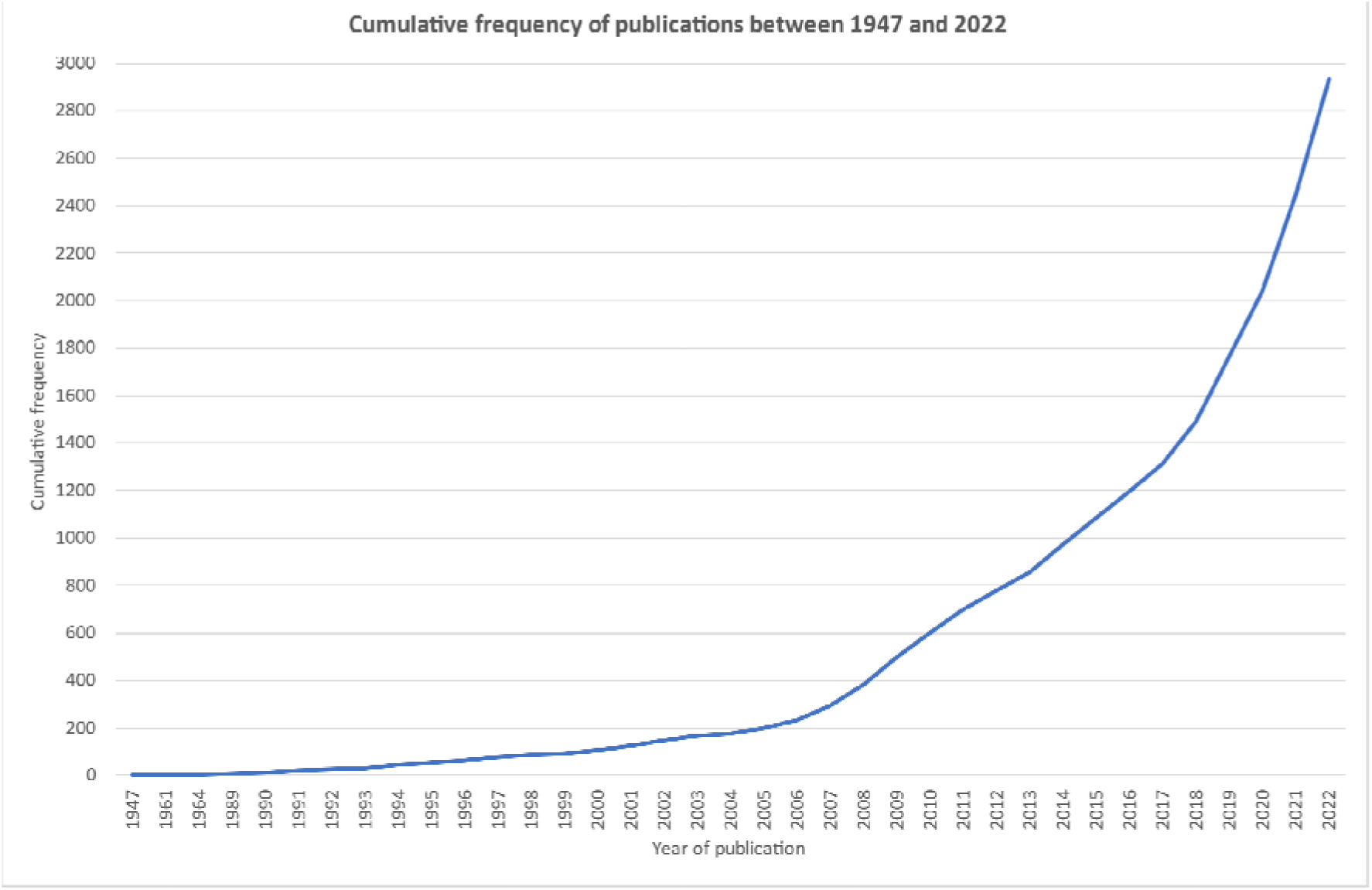
Cumulative frequency of articles published between 1947 and 2022 (n=2932)

The figures for 2021 and 2022 were inflated by the publication of the two joint editorials[11,12] across multiple journals. A total of 43 journals in our sample published the first editorial, 92 the second, and 23 both. Figure 4 shows the number of journals publishing articles on climate change for the first time; there was a big increase between 2017 and 2022.

**Figure 4:**
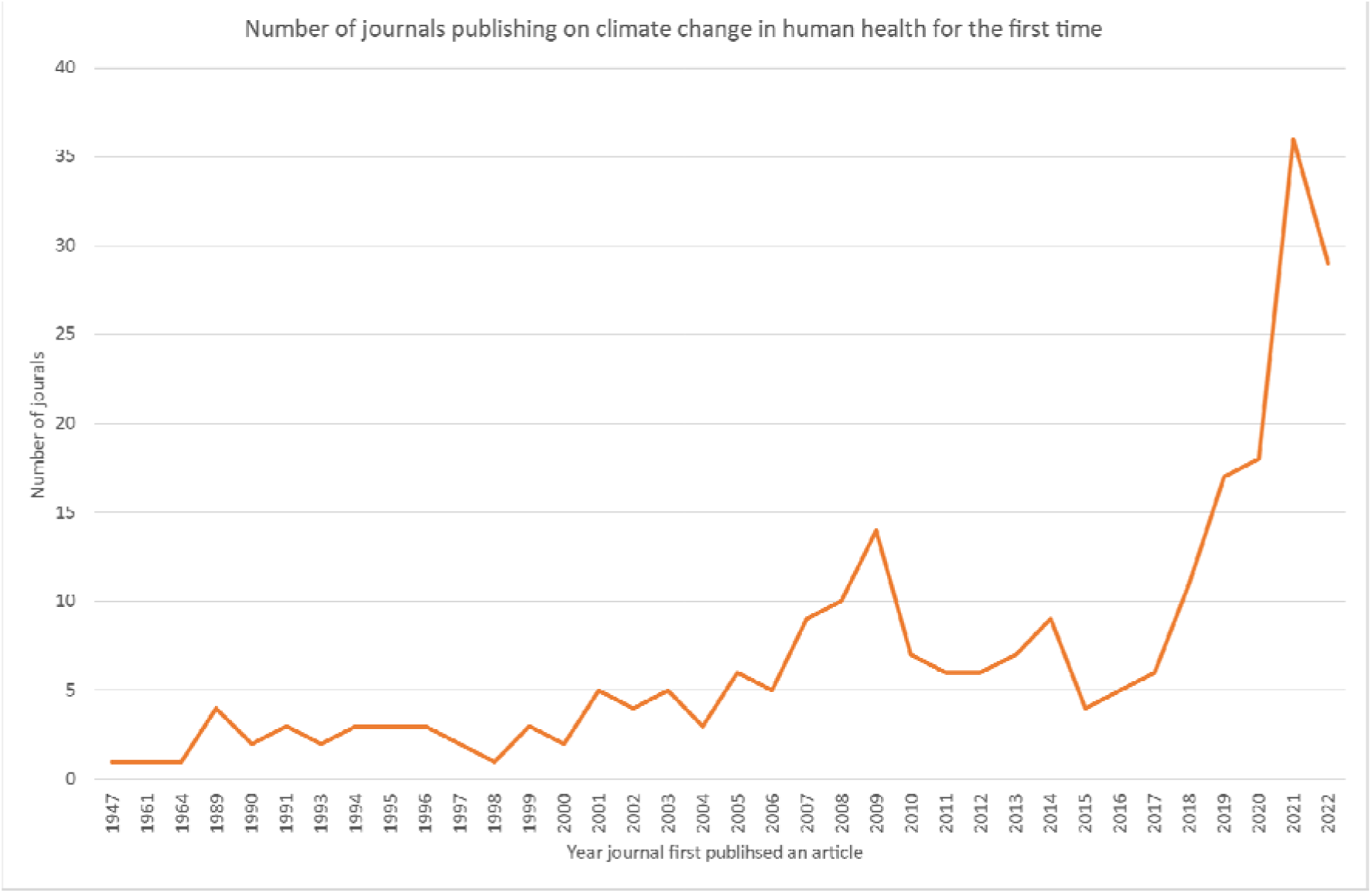
Number of journals publishing an article on climate change in human health for the first time (n=253 journals)

### Distribution across journals

The 2932 eligible articles were published in 253 of the 330 included journals between 1947 and 2022. While the articles were published in journals in 39 countries, two thirds (1929; 66%) of the articles were published in a journal published in the UK or the US (Supplementary Table 1). Figure 5 shows the distribution of articles across these 330 journals. Almost a quarter (77/330; 23%) of the journals published no articles on climate change and health, and almost three quarters (241/330; 73%) published five or fewer articles. Nine of the top 48 most highly cited clinical medicine journals (based on JCR 2021 and indexed in Medline) published no articles; 58 of the 77 journals that published no articles were indexed in the JCR 2021 category of “Medicine, General & Internal”.

**Figure 5:**
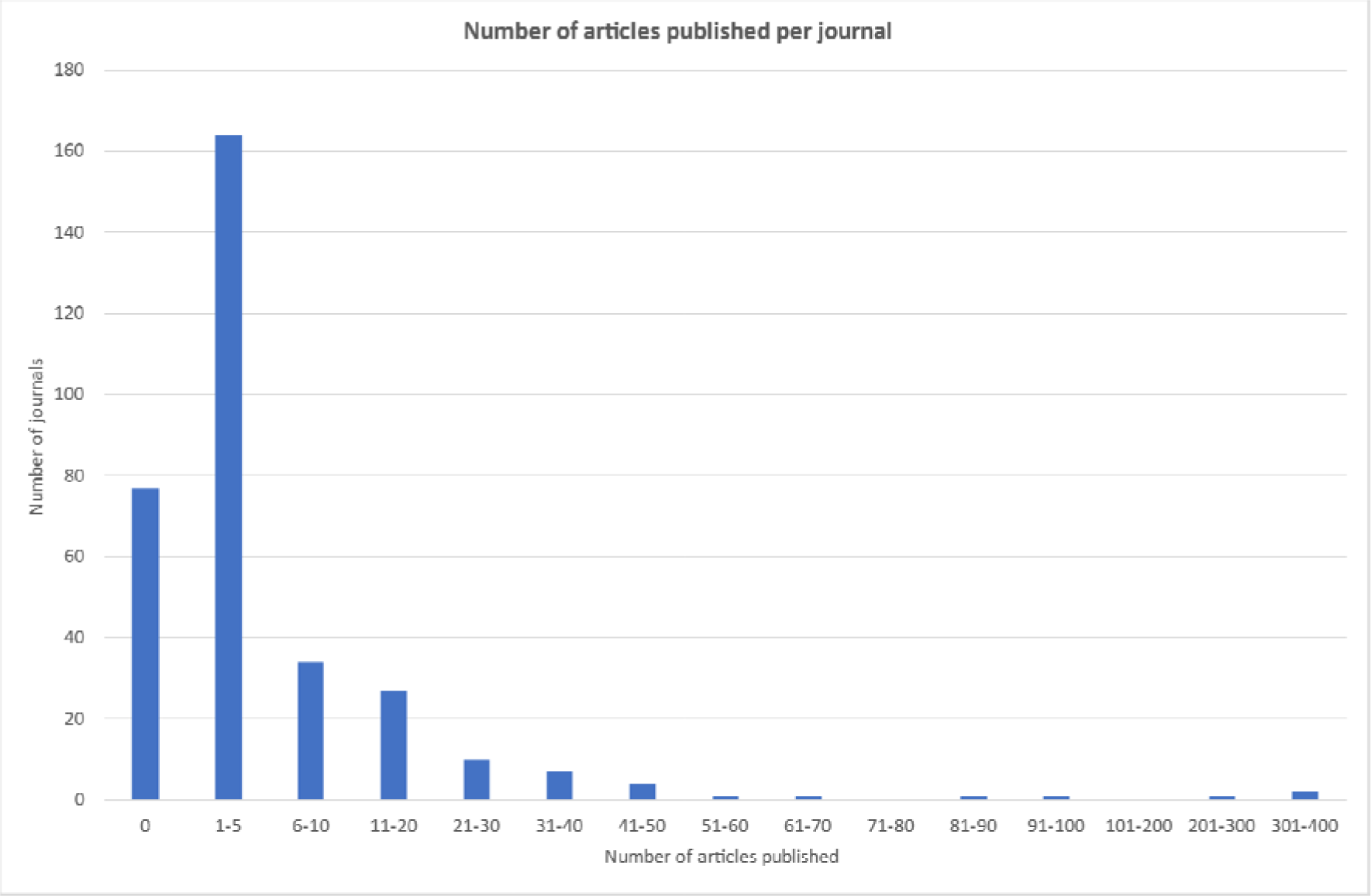
Distribution of articles across the 330 sampled journals between 1947 and 2022 (n=2932)

The ten journals publishing the most articles were *The BMJ* (343), *Lancet Planetary Health* (324), the *Lancet* (262), *Global Health Action* (92), the *Medical Journal of Australia* (82), *mBio* (67), *Bulletin of the World Health Organization* (55), *BMJ Open* (49), *JAMA* (44), and *CMAJ* (42). These 10 journals published 1360 (46%) of the 2932 articles.

Supplementary Table 2 shows the 59 journals publishing at least 10 articles between 1947 and 2022. Supplementary Figure 1 shows the number of articles published each year over time for the ten journals publishing the most content between 1947 to 2022 (1360 articles). Since 2018 the specialist journal *Lancet Planetary Health* has published the most per year and in 2021-2022 it published 153 (17%) of all the 895 articles published; *The BMJ* published 98 (11%) and the Lancet 37 (4%).

Supplementary Figure 2 shows the number of journals that published at least one article on climate change in human health between 1947 and 2022. In 2000 fewer than ten journals had published at least one article during the year; in 2010 it was slightly fewer than 40; in 2020 it was almost 100, and in 2022 it was 161 journals.

### Article characteristics

#### Language of publication

The 2932 articles were published in 10 languages (English 2795, Spanish 41, Norwegian 41,

French 31, German 22, Portuguese 19, Russian 6, Hungarian 5, Icelandic 3 and Chinese 1). 2795/2932 (95%) of the articles were published in English.

#### Article type

The publication type variable gave 134 different article types and we were not able to distinguish research from editorial and news type content without access to the full text of all articles.

However, 1345/2932 (46%) articles had an abstract (abstracts were introduced to MEDLINE in the 1960s) so it is likely that fewer than half were research papers.

#### Impact of the first joint editorial

The aim of the joint editorials published in multiple journals is to push governments to be more active in acting on climate change and to reach more health professionals with information on the importance of acting on climate change for the benefit of health. The first joint editorial[11] was published in September 2021 and the second[12] in October 2022, and these boosted the number of articles published on the topic and the number of journals publishing on the topic in those years. If we exclude the publications of both these joint editorials there were 2796 articles published across 224 journals; 106/330 (32%) journals would have published no articles on climate change between 1860 and 2022.

Of the 330 journals, 112 (34%) published at least one of the joint editorials and 32 (29%) of these 112 were publishing on the topic for the first time. 29 (26%) of the 112 publishing at least one of the editorials did not publish anything else on the topic between 1947 and 2022. Of the 23 journals that published both of the editorials, 5 (22%) did not publish anything else on the topic between 1947 and 2022.

A total of 43 (13%) of the 330 journals published the first joint editorial. While 30 (70%) of these 43 journals published at least one article on climate change after this first joint editorial, for 10/30 (33%) this was just the second joint editorial.

18 (42%) of the 43 journals that published the first joint editorial had not previously published any articles on climate change in human health; 8/18 (44%) journals went on to publish at least one more article on the topic but 6 of these 8 just published the second joint editorial.

## DISCUSSION

### Main findings

Our study confirms that there were occasional articles published on climate change and human health in the 1990s, but the increase in the number of articles began in the early 2000s.

Our study looked at journals as well as articles, and the main finding that our study adds to previous studies is that despite climate change being recognised as the major threat to global health, roughly a quarter of health journals (77/330; 23%) had by 2022 published nothing on the topic and almost three quarters (241/330; 73%) had published five or fewer articles. If the joint editorials published in multiple journals are excluded from the analysis then almost a third (106/330; 32%) of the journals would have published no articles on climate change between 1860 and 2022.

Three journals (*The BMJ*, *Lancet Planetary Health,* and *Lancet*), all based in Britain, published almost a third of the articles (32%), and ten journals, all in English, published almost half (46%). The two leading general journals based in Britain (*Lancet* and *The BMJ*) published 605 (21%) articles between 1860 and 2022, while the two leading general journals based in the US (*NEJM* and *JAMA*) published 83 (3%).

Another new finding is that the joint editorials published in multiple journals in 2021 and 2022 were often the first article on climate change and health that the journals published: 112 of the 330 journals published at least one of the joint editorials and 32 (29%) were publishing on the topic for the first time.

Publishing the joint editorial as their first article on climate change has so far had little impact on encouraging the journals to publish more: of the 18 journals that published the first joint editorial as their first article on climate change, fewer than half (8, 44%) published at least one more article on the topic and six of these eight just published the second joint editorial.

### Our results in comparison to other studies

Studies that have examined articles published on climate change and health have shown that articles first appeared in the early 1990s and then rapidly increased from the early 2000s.[3, 8, 7, 10, 13, 15, 16, 18] One study found, however, that papers concerned with climate change are published at about twice the rate in other sectors, when compared with health.[8]

The only study that gives information on journals is that by Sangam and Savitha, which looked at all articles on climate change in Web of Science between 2001 and 2016.[15] The study includes data on the 20 journals publishing the most publications: *Science* is at the top (1523 publications). None of the 20 were health journals.

Other studies found that most articles on climate change and health came from high income countries. We didn’t look at the country of origin of the articles, but we did find that most articles were published in journals from high income countries. Verner *et al.* found that about two-thirds of all published studies were carried out in OECD (Organization for Economic Cooperation and Development) countries, predominantly in Europe and North America.[8] Berrang Ford *et al.* looked at 15,963 studies published between 2013 and 2019 and found the studies came mostly from high-income countries and China. There were few studies from low-income countries despite them suffering most from the health consequences of climate change.[7] Bartlett *et al.* studied research articles between 2012 and 2019, took a random sample of 348, and found that fewer than a third had first authors from the Global South.[10] The Lancet Countdown looked at scientific studies and the majority of studies were located in, and led by, authors in WHO (World Health Organisation) regions of Western Pacific and the Americas.[3]

We did not analyse the subject matter of the articles that we studied, but others have. Articles on the health impacts of climate change far outnumbered those on mitigation and adaptation, with little or nothing on decarbonising health systems.[3] Berrang Ford *et al.* found that air quality and heat stress were the most frequently studied exposures, with major gaps in evidence on the impact of climate change on mental health, undernutrition, and maternal and child health.[7] Bartlett *et al.* found a lack of research on vulnerable populations, particularly the elderly and those in the Global South.[10] Verner *et al.* found that the effects of climate change on malnutrition and non-communicable diseases is understudied.[8]

### Study strengths and limitations

A strength of our study is that we have looked at journals not just articles. We could not find any other studies that had looked at journals rather than articles, although Sangam and Savitha gave data on the top 20 journals covering all aspects of the science of climate not just health.[15] We were able to examine the distribution of articles in journals and how many had published little or nothing on climate change and health.

We chose to use Ovid MEDLINE as the database to search in order to use complex adjacent word searching, which was not possible in PubMed. We may have missed some articles, but were able to design a more specific search using this technique. In addition to keyword searches, we also used MeSH headings, however the MeSH heading “Climate Change” is relatively new and terminology has changed over time. Thus, we used combinations of specific MeSH headings (e.g., Climate Change, Global Warming) that were directly related to our topic and MeSH headings that were more generic (e.g., Public Health, Global Health) in combination with keyword searches.

Our use of search terms and adjacent word searching to try to capture all relevant papers will inevitably have resulted in the inclusion of some articles that may not be devoted to climate change’s impact on human health and may just have mentioned it. There is also an element of subjectivity when determining if an article is *about* climate change and human health or just *mentioning* it. Therefore, to improve the accuracy of the search results, two independent raters screened the articles’ title and abstract (where available) for article eligibility. Checking the full text of the included articles for relevance was not feasible as we were unable to access a large number of articles due to subscription and language barriers.

This independent screening of the MEDLINE search results is a strength in our approach, as other studies did not include this step, with the exception of Bartlett *et al.,*[10] and they studied only articles from North America published after 2012. However, even screening didn’t remove all ineligible articles. For example, we included all articles that had “climate change” in the title but closer inspection of the earliest article, “*Dangers of climate change*”, published in German in *Schweizerische Medizinische Wochenschrift* in 1947[19] showed that it was about high altitude acclimatisation. A further two articles were published before 1989 (both in the 1960s) and are published in Spanish[20] and French[21] with no abstract. These articles may also not have been on the topic of climate change as they were published before the adoption of the use of the term climate change as we use it today. We expect that these errors will only have impacted the very early publications, but for reasons of consistency have not excluded them from the analysis as we did not apply full text screening across the whole sample.

Extreme weather events - heatwaves, floods, wildfires, and extreme storms - have become more common with climate change, but existed before climate change and are not always linked to climate change. We reviewed a sample of articles on extreme weather events and found that only 2% mentioned these events being linked to climate change. Other search strategies might lead to these articles being included.

Our study includes the intervention of a group organising a joint editorial on climate change and health, which was published in multiple journals in 2021 and 2022. We were able to examine whether journals that published the first editorial, not having published before on climate change and health, were prompted to publish more articles on climate change and health.

We must be cautious in generalising to all health journals from our study. It was difficult to identify a representative list of health journals to include and our sampling will inevitably have influenced the results. We did include the most highly cited clinical journals and the top journals in general and internal medicine, which means that we have a bias towards bigger journals and medical as opposed to health journals. These journals are better resourced than most health journals, which makes it likely that we probably underestimated the number of health journals that have never mentioned climate change.

We may also have excluded some relevant articles. For example, by including only journals that are currently indexed in MEDLINE, we excluded many journals that published the joint editorials on climate change.[11, 12] However, we needed to do the article searches electronically as hand searching of the entire back archive of 330 journals was not feasible. Not all articles in a journal are always indexed in MEDLINE even if the journal itself is indexed. Some journals are indexed with multiple titles in MEDLINE and there is no systematic way of identifying these, so some journal names may have been omitted.

We excluded all the results from our extreme weather-related search, as our assessment of a random sample of 5% of these articles indicated that only 2% were potentially eligible for inclusion. However, we know that we missed a very small proportion of articles by excluding these results.

A limitation of our study is that we have not been able to examine the types of articles published, so our study does not differentiate between research articles and editorials, comments, news, letters, and other types of articles. We know that only 46% of the articles had abstracts, which suggests that fewer than half of the articles were research studies. We know too that funding for research on climate change and health has been slow to become available,[22] meaning that early articles - those before 2010 - were likely to be editorials and comments rather than research. This may also explain why particular journals have published more--for instance, both the Lancet and *The BMJ* publishes news.

Nor do we know the themes of the articles, but we know from the Lancet Countdown[3] that the majority of articles published have been about the impact of climate change on health, far fewer on mitigating the effects of climate change, and fewer still on adapting to the changes that have already happened. Almost no articles so far have been on decarbonising health systems.

Another limitation of the study may have been unconscious bias as several of the authors were linked with the BMJ and the UK Health Alliance on Climate Change (see Competing Interest statement).

### Study implications

#### Why have most health journals published little or nothing on climate change and health?

Despite climate change being predicted in the 19th century, detectable effects occurring by the ‘80s, and the first Earth Summit being held in 1992, few articles on climate change and health appeared in health journals until the 2000s. Indeed, despite climate change being identified as *the* major threat to global health, by 2022 about a quarter of health journals in our sample had published nothing on climate change and health and almost three quarters had published five or fewer articles.

Why have health journals been so slow? There are almost certainly multiple reasons.

One answer might be that almost everybody has been slow. Although the serious threat of climate change has long been identified, emissions of greenhouse gases are still rising.[3] The annual UN meeting on climate change, COP, will feature health in the main agenda for the first time only in 2023. Of course, the policymakers may have been slow because health scientists have been slow.

Another reason may be that journals found the subject of climate change “too political.”[23] Although scientists have been clear since the ‘90s that climate change is damaging health, many politicians denied the existence of climate change, that it is caused by human activity, and that it matters. Some still do. This may have made editors of health journals nervous about covering the subject. That journals based in the US, where climate change has been a party political issue, have been slower than journals based in Britain, where there has been consensus on the importance of climate change, offers support for this possible reason.

Although general, public and global health, and epidemiology journals could feel comfortable covering climate change and health, specialist journals may have worried that the topic was not relevant to their readers. The health community has now recognised that health systems have to decarbonise, which means that every specialty will have to explore how it can do so, making the subject directly relevant to all parts of healthcare.

More than half of the articles we identified are not research articles but editorials, news, letters, comments, and educational articles. Many health journals publish little but research articles, and, as we and others have shown, there has been little research on climate change and health until recently.

One important reason for the slow and limited coverage of climate change and health is that many journals commission few articles. They wait for material, usually research papers, to be submitted. A related reason is that even if they do commission material, many health editors would not know whom to ask to write on the subject: there have been, and still are, few authorities on climate change and health.

#### Increasing coverage of climate change and health in health journals?

Coverage of climate change and health is increasing in health journals and is likely to continue to increase because of growing global concern about the seriousness of the climate crisis, and because of the recognition of the need for decarbonising health care.

It seems that many journals are willing to publish further articles on climate change and health but lack the capacity to commission such articles. This finding supports a proposal to establish a consortium that would lead the publishing of joint editorials on the subject, perhaps more than once a year, and commission and share articles with the many journals that lack the capacity to commission articles themselves. In this way, material on climate change and health could reach many more health professionals.

## Author contributions

SS, JM, MLR, BR, RS designed the study and developed the search strategy. MLR wrote the search strategy and conducted the literature searches. SS, MLR, and JM extracted data. RS, BR, JM, FW, AP, MLR screened the articles for eligibility. SS analysed the data with JM. SS is the study guarantor. All authors had direct access to the data, interpreted the findings, and contributed to the writing of the manuscript. The corresponding author attests that all listed authors meet authorship criteria and that no others meeting the criteria have been omitted.

## Funding

No specific funding.

## Competing interests

All authors have completed the ICMJE uniform disclosure form at www.icmje.org/disclosure-of-interest/ and declare: no support from any organisation for the submitted work; SS is a full time employee of BMJ Publishing Group which published many of the articles included in this study. MLR is working on a self-funded PhD in conjunction with BMJ Publishing Group and Maastricht University. AP is working as a policy officer with the UK Health Alliance on Climate Change (UKHACC). RS was the editor of the *BMJ* from 1991-2004, an assistant editor before that from 1979, and is now chair of the UK Health Alliance on Climate Change, which organised and funded the joint editorial that has appeared in multiple journals. RS has long been concerned about environmental issues. FW recently completed a year as a sustainability fellowship at *The BMJ*, and she remains a freelance clinical editor at *The BMJ*. BR worked at UKHACC for two months in autumn 2022 and supported work on the second joint editorial. JM helped with the second joint editorial.

## Ethical approval

Not required.

## Data availability statement

The data that support the findings of this study are available on request.

---

**Supplementary Table 1:** Journals’ country of publication on the date articles were published.

| **Country** | **Number of articles published** | **% of all articles published** |
| --- | --- | --- |
| England | 1319 | 45% |
| United States | 610 | 21% |
| Netherlands | 375 | 13% |
| Australia | 139 | 5% |
| Switzerland | 64 | 2% |
| Canada | 58 | 2% |
| China | 43 | 1% |
| Norway | 41 | 1% |
| New Zealand | 37 | 1% |
| Germany | 29 | 1% |
| Iran | 25 | 1% |
| Brazil | 23 | 1% |
| Denmark | 22 | 1% |
| India | 19 | 1% |
| Spain | 19 | 1% |
| Austria | 13 | 0% |
| Chile | 13 | 0% |
| France | 11 | 0% |
| Russia (Federation) | 8 | 0% |
| Argentina | 7 | 0% |
| Singapore | 7 | 0% |
| Pakistan | 6 | 0% |
| Portugal | 6 | 0% |
| Hungary | 5 | 0% |
| South Africa | 5 | 0% |
| Korea (South) | 4 | 0% |
| Turkey | 4 | 0% |
| Iceland | 3 | 0% |
| Uganda | 3 | 0% |
| Ireland | 2 | 0% |
| Japan | 2 | 0% |
| Mexico | 2 | 0% |
| Saudi Arabia | 2 | 0% |
| Bangladesh | 1 | 0% |
| Bosnia and Herzegovina | 1 | 0% |
| Israel | 1 | 0% |
| Italy | 1 | 0% |
| Poland | 1 | 0% |
| Slovakia | 1 | 0% |

Note - One journal was published in 2 different countries (England and Germany)

**Supplementary Table 2:**
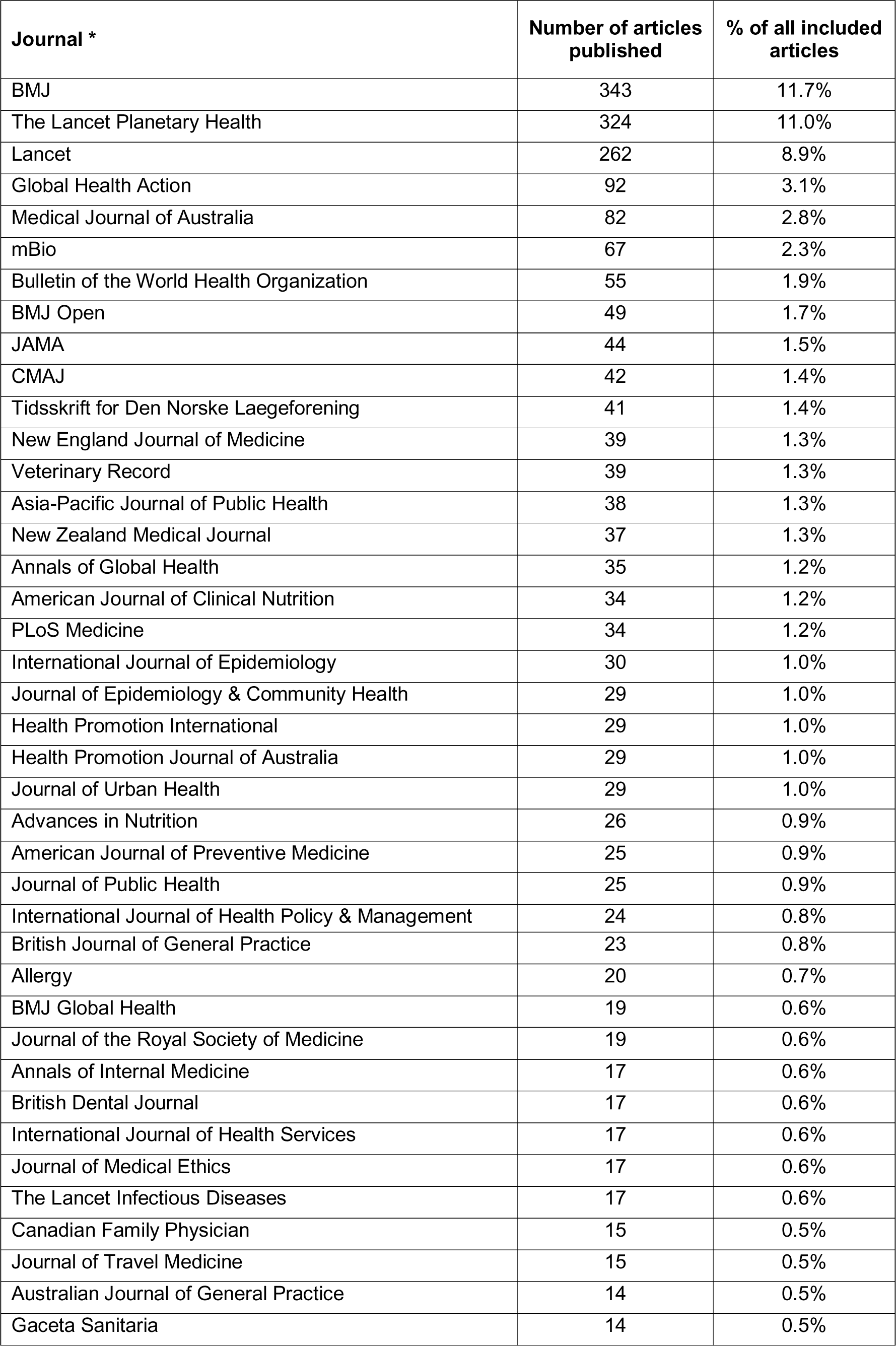
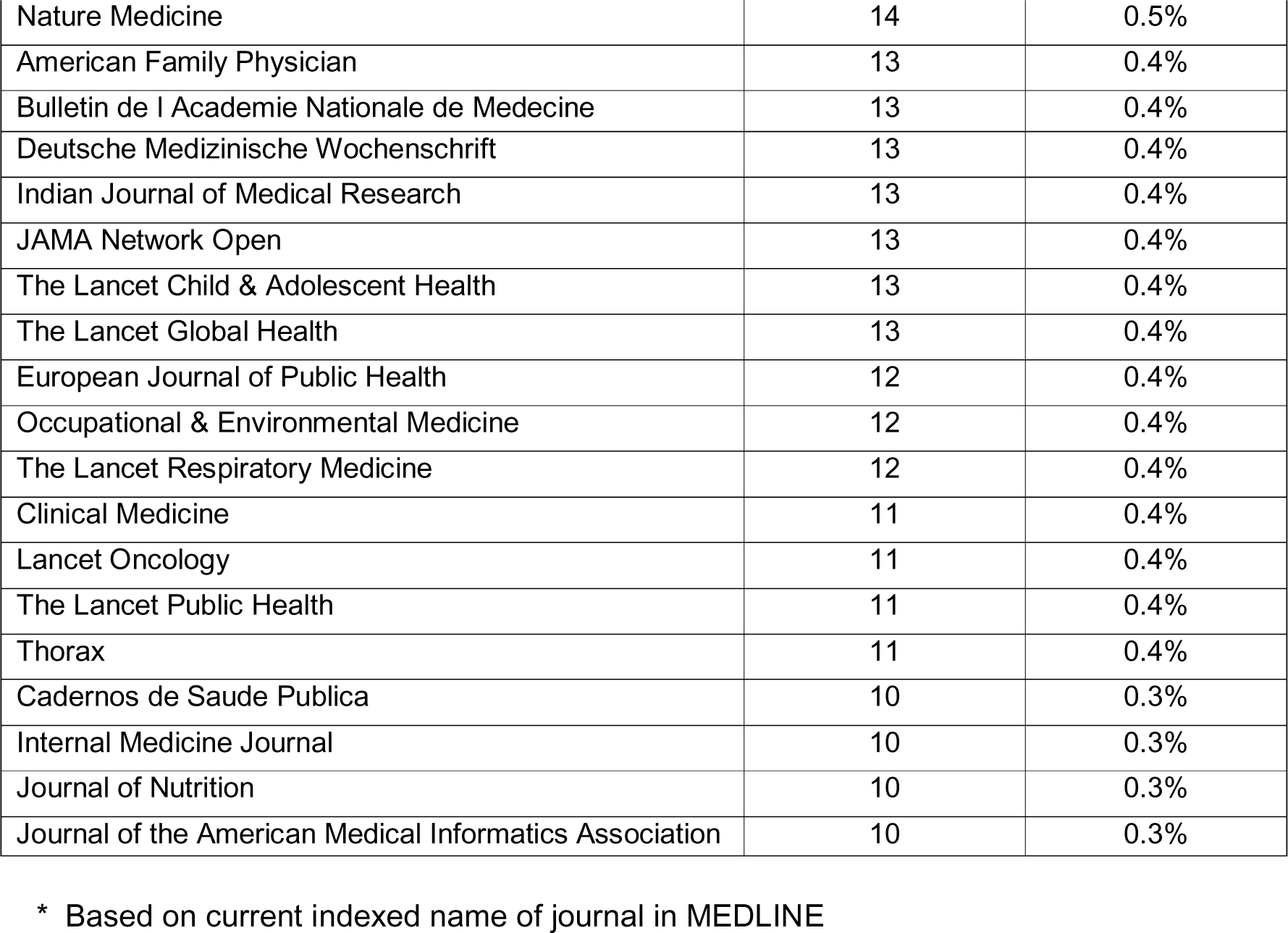
Journals publishing at least 10 articles on climate change in human health between 1947 and 2022.

**Supplementary Figure 1:**
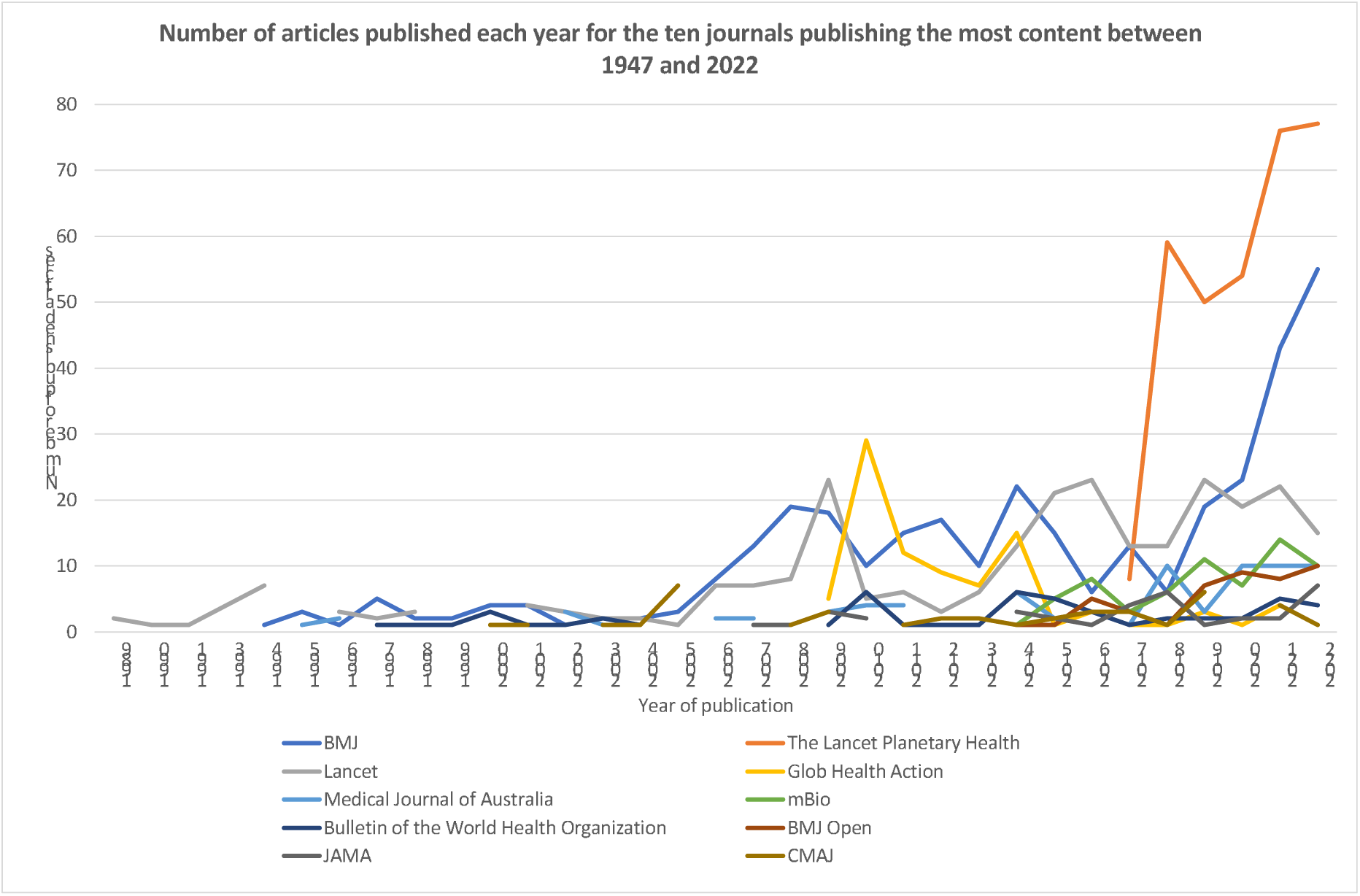
Number of articles published each year for the top ten journals publishing the most content between 1947 and 2022.

**Supplementary Figure 2:**
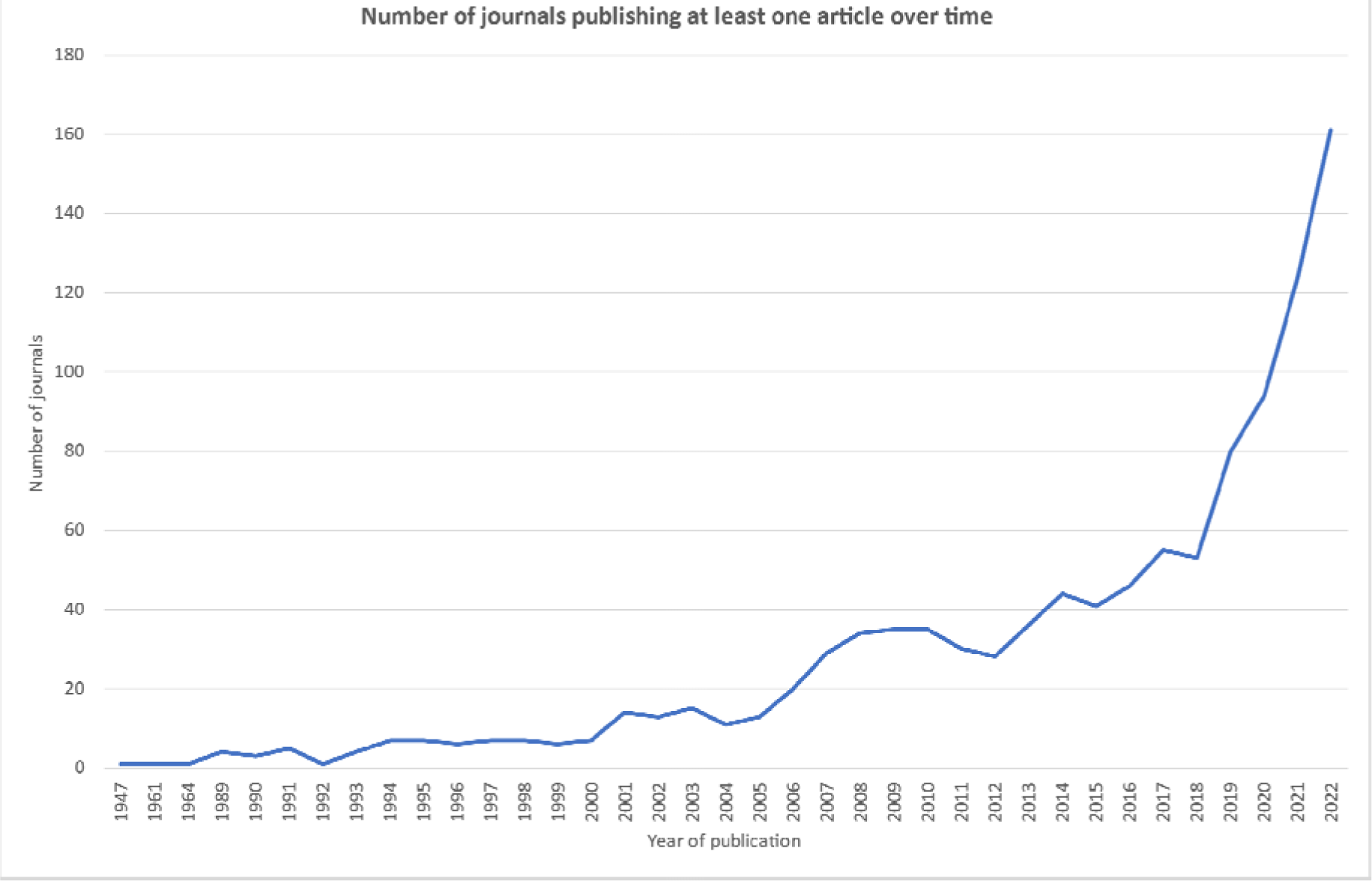
Number of journals publishing at least one article each year between 1947 and 2022 (n=330)

### Appendix 1 Search strategy

Search Date: 2023-01-26

Line 409 is the set of results for the main search used in this study.

Line 410 is the set of results for the extreme weather condition articles that are potentially eligible

Ovid MEDLINE(R) ALL <1946 to January 25, 2023>

1 ((climat* or planetary) adj3 (disaster* or warm* or chang* or sustainab* or crisis or crises or justice* or security or emergenc* or action* or impact* or anthropogen* or induced or extrem* or catastroph* or breakdown* or vulnerab* or resilien* or anxiety or tipping point* or activis* or pact)).ti,ab,kf. 72203

2 planetary health.ti,ab,kf. 851

3 (warming adj6 (carbon or ocean* or ice or lake* or stratospher* or atmospher* or global* or ecosytem*)).ti,ab,kf. 14789

4 (greenhouse adj3 (gas* or emission* or effect* or warm*)).ti,ab,kf. 14499

5 (climat* or weather or planetary).ti,ab. 158545

6 exp Biodiversity/ 122075

7 exp Public Health/ 9072852

8 exp Global Health/ 54707

9 6 or 7 or 8 9170217

10 5 and 9 59378

11 exp climatic processes/ 71289

12 exp extreme weather/ 276

13 exp Extreme Heat/ 537

14 exp ozone depletion/ 65

15 exp sea level rise/ 121

16 exp stratospheric ozone/ 104

17 exp Temperature/ 447416

18 11 or 12 or 13 or 14 or 15 or 16 or 17 506630

19 9 and 18 124268

20 exp Greenhouse Gases/ or exp Greenhouse Effect/ 7747

21 exp Global Warming/ 4242

22 exp Climate Change/ 28643

23 exp Anthropogenic Effects/ 393

24 anthropogen*.ti,ab,kf. 35903

25 anthropocene.ti,ab,kf. 1143

26 (cop27 or cop26 or cop 25 or cop24 or cop23 or cop22 or cop21 or cop20 or cop19 or cop18 or cop17 or cop16 or cop15 or cop14 or cop13 or cop12 or cop11 or cop10 or cop9 or cop8 or cop7 or cop6 or cop5 or cop4 or cop3 or cop2).ti,ab. 1252

27 (kyoto protocol or paris agreement).ti,ab,kf. 736

28 (climat* or planetary or greenhouse or global warm*).ti,ab,kf. 177664

29 (human* or health*).ti,ab. 6124267

30 (extreme adj1 (weather or heat or cold)).ti,ab. 3693

31 28 or 29 6260882

32 30 and 31 2554

33 (hurricane* or drought* or famine* or food scarcity or flood* or landslide* or heat wave* or heatwave* or mass extinct* or typhoon* or ((tropic* or dust or extreme or catastroph* or danger*) and storm*)).ti,ab. 63983

34 ((carbon or methane*) adj2 (sink* or sequestrat* or budget* or footprint* or emission*)).ti,ab. 16413

35 deforestation.ti,ab. 3340

36 (zero adj1 (emission* or net)).ti,ab. 1116

37 (eco-anxi* or ecoanxi*).ti,ab,kf. 59

38 (acid* adj3 (rain* or ocean*)).ti,ab. 3842

39 (crop or crops or soil).ti,ab. 263615

40 biodivers*.ti,ab,kf. 38830

41 33 or 34 or 35 or 36 or 37 or 38 or 39 or 40 364397

42 28 and 41 50854

43 1 or 2 or 3 or 4 or 10 or 19 or 20 or 21 or 22 or 23 or 24 or 25 or 26 or 27 or 32 or 42 274507

44 ((climat* or planetary) adj3 (disaster* or warm* or chang* or sustainab* or crisis or crises or justice* or security or emergenc* or action* or impact* or anthropogen* or induced or extrem* or catastroph* or breakdown* or vulnerab* or resilien* or anxiety or tipping point* or activis* or pact)).ti,ab,kf. 72203

45 planetary health.ti,ab,kf. 851

46 (warming adj6 (carbon or ocean* or ice or lake* or stratospher* or atmospher* or global* or ecosytem*)).ti,ab,kf. 14789

47 (greenhouse adj3 (gas* or emission* or effect* or warm*)).ti,ab,kf. 14499

48 (climat* or planetary).ti,ab. 140435

49 exp Biodiversity/ 122075

50 exp Global Health/ 54707

51 49 or 50 176604

52 48 and 51 8235

53 exp sea level rise/ 121

54 48 and 53 61

55 exp Greenhouse Gases/ or exp Greenhouse Effect/ 7747

56 exp Global Warming/ 4242

57 exp Climate Change/ 28643

58 anthropogen*.ti,ab,kf. 35903

59 anthropocene.ti,ab,kf. 1143

60 (cop27 or cop26 or cop 25 or cop24 or cop23 or cop22 or cop21 or cop20 or cop19 or cop18 or cop17 or cop16 or cop15 or cop14 or cop13 or cop12 or cop11 or cop10 or cop9 or cop8 or cop7 or cop6 or cop5 or cop4 or cop3 or cop2).ti,ab. 1252

61 (kyoto protocol or paris agreement).ti,ab,kf. 736

62 (climat* or planetary or greenhouse or global warm*).ti,ab,kf. 177664

63 (hurricane* or drought* or famine* or food scarcity or flood* or landslide* or heat wave* or heatwave* or mass extinct* or typhoon* or ((tropic* or dust or extreme or catastroph* or danger*) and storm*)).ti,ab. 63983

64 ((carbon or methane*) adj2 (sink* or sequestrat* or budget* or footprint* or emission*)).ti,ab. 16413

65 deforestation.ti,ab. 3340

66 (zero adj1 (emission* or net)).ti,ab. 1116

67 (eco-anxi* or ecoanxi*).ti,ab,kf. 59

68 (acid* adj3 (rain* or ocean*)).ti,ab. 3842

69 (crop or crops or soil).ti,ab. 263615

70 biodivers*.ti,ab,kf. 38830

71 63 or 64 or 65 or 66 or 67 or 68 or 69 or 70 364397

72 62 and 71 50854

73 44 or 45 or 46 or 47 or 52 or 54 or 55 or 56 or 57 or 58 or 59 or 60 or 61 or 72 147854

74 43 not 73 126653

75 ca a cancer journal for clinicians.jn. 2405

76 lancet.jn. 141576

77 new england journal of medicine.jn. 83452

78 (jama or journal of the american medical association).jn. 89642

79 nature reviews immunology.jn. 2813

80 lancet respiratory medicine.jn. 3069

81 (bmj or british medical journal or british medical journal clinical research ed).jn. 199208

82 nature medicine.jn. 11564

83 lancet microbe.jn. 539

84 lancet psychiatry.jn. 2645

85 nature reviews gastroenterology & hepatology.jn. 2399

86 lancet public health.jn. 1023

87 lancet infectious diseases.jn. 6974

88 nature reviews cancer.jn. 2473

89 nature reviews disease primers.jn. 690

90 nature reviews clinical oncology.jn. 2499

91 lancet neurology.jn. 4935

92 nature reviews genetics.jn. 2682

93 lancet oncology.jn. 8803

94 annals of oncology.jn. 12563

95 annals of internal medicine.jn. 36439

96 journal of clinical oncology.jn. 26167

97 nature reviews cardiology.jn. 2640

98 nature reviews endocrinology.jn. 2584

99 lancet gastroenterology & hepatology.jn. 1359

100 lancet diabetes & endocrinology.jn. 2146

101 nature reviews neurology.jn. 2363

102 (jama internal medicine or archives of internal medicine or “ama archives of internal medicine“).jn. 26961

103 immunity.jn. 5674

104 nature reviews nephrology.jn. 2316

105 (intensive care medicine or “european journal of intensive care medicine“).jn. 12184

106 nature genetics.jn. 8820

107 circulation.jn. 46316

108 journal of travel medicine.jn. 2810

109 lancet global health.jn. 3076

110 nature reviews neuroscience.jn. 2677

111 journal of infection.jn. 7472

112 cancer cell.jn. 3424

113 cancer discovery.jn. 3823

114 lancet child & adolescent health.jn. 1014

115 lancet digital health.jn. 527

116 morbidity & mortality weekly report recommendations & reports.jn. 430

117 european heart journal.jn. 21264

118 mmwr morbidity & mortality weekly report.jn. 9551

119 military medical research.jn. 430

120 gastroenterology.jn. 32460

121 european respiratory journal.jn. 15188

122 jama oncology.jn. 3450

123 journal of the royal society of medicine.jn. 11418

124 (canadian medical association journal or cmaj canadian medical association journal).jn. 56623

125 (qjm or quarterly journal of medicine).jn. 7778

126 jama network open.jn. 7809

127 (journal of internal medicine or acta medica scandinavica).jn. 12391

128 medical journal of australia.jn. 44074

129 “journal of cachexia sarcopenia and muscle“.jn. 1241

130 cochrane database of systematic reviews.jn. 16112

131 plos medicine public library of science.jn. 4683

132 bmc medicine.jn. 3626

133 mayo clinic proceedings.jn. 11472

134 (translational research the journal of laboratory & clinical medicine or journal of laboratory & clinical medicine).jn. 14109

135 annals of the academy of medicine singapore.jn. 6635

136 deutsches arzteblatt international.jn. 4187

137 european journal of internal medicine.jn. 4911

138 medical clinics of north america.jn. 7421

139 american journal of preventive medicine.jn. 7593

140 amyloid.jn. 1375

141 cleveland clinic journal of medicine.jn. 4501

142 journal of general internal medicine.jn. 11197

143 (british journal of general practice or journal of the royal college of general practitioners).jn. 13235

144 journal of evidence based medicine.jn. 553

145 chinese medical journal.jn. 18951

146 (american journal of chinese medicine or comparative medicine east & west).jn. 2981

147 american journal of medicine.jn. 26820

148 british medical bulletin.jn. 3779

149 (journal of urban health or bulletin of the new york academy of medicine).jn. 7966

150 european journal of clinical investigation.jn. 6632

151 palliative medicine.jn. 3119

152 annals of family medicine.jn. 2213

153 minerva medica.jn. 26160

154 journal of pain & symptom management.jn. 6417

155 internal & emergency medicine.jn. 3151

156 clinical medicine.jn. 5000

157 journal of korean medical science.jn. 7281

158 annals of medicine.jn. 3104

159 (american family physician or american family physician gp).jn. 13875

160 indian journal of medical research.jn. 14149

161 panminerva medica.jn. 3590

162 (polish archives of internal medicine or polskie archiwum medycyny wewnetrznej).jn. 10407

163 postgraduate medical journal.jn. 17094

164 (bmj evidence based medicine or evidence based medicine).jn. 2289

165 preventive medicine.jn. 8535

166 disease a month.jn. 1485

167 postgraduate medicine.jn. 15579

168 (swiss medical weekly or schweizerische medizinische wochenschrift journal suisse de medecine).jn. 22306

169 (journal of the formosan medical association or taiwan i hsueh hui tsa chih journal of the formosan medical association).jn. 9098

170 international journal of medical sciences.jn. 2728

171 pain medicine.jn. 5040

172 european journal of general practice.jn. 850

173 (international journal of evidence based healthcare or jbi evidence implementation).jn. 612

174 balkan medical journal.jn. 1045

175 american journal of the medical sciences.jn. 11801

176 (“journal of the chinese medical association jcma” or chung hua i hsueh tsa chih chinese medical journal).jn. 22959

177 singapore medical journal.jn. 8382

178 american journal of managed care.jn. 4600

179 (journal of investigative medicine or clinical research).jn. 2438

180 medicina clinica.jn. 22217

181 korean journal of internal medicine.jn. 2815

182 international journal of clinical practice.jn. 6767

183 scandinavian journal of primary health care.jn. 1984

184 archives of iranian medicine.jn. 2329

185 systematic reviews.jn. 2497

186 revista clinica espanola.jn. 18438

187 yonsei medical journal.jn. 5100

188 canadian family physician.jn. 12934

189 journal of womens health.jn. 4060

190 bmj open.jn. 29275

191 journal of medical economics.jn. 1905

192 (medicina kaunas lithuania or medicina kaunas).jn. 7305

193 Turkish journal of medical sciences.jn. 2337

194 journal of hospital medicine online.jn. 2767

195 (clinics sao paulo brazil or “revista do hospital das clinicas faculdade de medicina da universidade de sao paulo“).jn. 5792

196 (bmj military health or journal of the royal army medical corps).jn. 3911

197 journal of the national medical association.jn. 11673

198 current medical research & opinion.jn. 6918

199 revista de investigacion clinica.jn. 4005

200 (upsala journal of medical sciences or acta societatis medicorum upsaliensis).jn. 2194

201 (bmc family practice or bmc primary care).jn. 2978

202 (internal medicine journal or “australian & new zealand journal of medicine“).jn. 9876

203 tohoku journal of experimental medicine.jn. 8880

204 scottish medical journal.jn. 4191

205 croatian medical journal.jn. 2424

206 (journal of the american board of family medicine jabfm or “journal of the american board of family practice“).jn. 3923

207 journal of evaluation in clinical practice.jn. 3551

208 family practice.jn. 3905

209 wiener klinische wochenschrift.jn. 16369

210 family medicine.jn. 5081

211 atencion primaria.jn. 7548

212 primary care clinics in office practice.jn. 2622

213 south african medical journal.jn. 849

214 medical principles & practice.jn. 2066

215 irish journal of medical science.jn. 7485

216 (australian journal of general practice or australian family physician).jn. 10113

217 (danish medical journal or danish medical bulletin).jn. 4127

218 (sao paulo medical journal revista paulista de medicina or “revista paulista de medicina“).jn. 5182

219 (presse medicale or nouvelle presse medicale).jn. 41451

220 medicine.jn. 34140

221 libyan journal of medicine.jn. 542

222 colombia medica.jn. 403

223 (revista da associacao medica brasileira or amb revista da associacao medica brasileira).jn. 6827

224 annals of saudi medicine.jn. 3872

225 acta medica portuguesa.jn. 4682

226 acta clinica belgica.jn. 4589

227 israel medical association journal imaj.jn. 5455

228 chronic illness.jn. 620

229 journal of postgraduate medicine.jn. 3961

230 bratislavske lekarske listy.jn. 9509

231 military medicine.jn. 15361

232 medizinische klinik intensivmedizin und notfallmedizin.jn. 1308

233 saudi medical journal.jn. 7238

234 (jaapa or journal of the american academy of physician assistants).jn. 3598

235 (british journal of hospital medicine or hospital medicine london).jn. 10856

236 (internal medicine or japanese journal of medicine).jn. 15532

237 hong kong medical journal.jn. 3341

238 nigerian journal of clinical practice.jn. 2971

239 (journal of nippon medical school nihon ika daigahu zasshi or journal of nippon medical school nihon ika daigaku zasshi or nippon ika daigaku zasshi journal of the nippon medical school).jn. 3163

240 african health sciences.jn. 2534

241 (netherlands journal of medicine or folia medica neerlandica).jn. 4718

242 jcpsp journal of the college of physicians & surgeons pakistan.jn. 6402

243 jpma journal of the pakistan medical association.jn. 11004

244 medicina.jn. 9467

245 acta clinica croatica.jn. 1364

246 revue de medecine interne.jn. 7215

247 (internist or innere medizin heidelberg germany).jn. 9597

248 southern medical journal.jn. 23343

249 medical problems of performing artists.jn. 469

250 journal of family practice.jn. 9676

251 gaceta medica de mexico.jn. 6741

252 orvosi hetilap.jn. 28568

253 revista medica de chile.jn. 13344

254 deutsche medizinische wochenschrift.jn. 37124

255 national medical journal of india.jn. 3736

256 jnma journal of the nepal medical association.jn. 1735

257 laeknabladid.jn. 1745

258 terapevticheskii arkhiv.jn. 20851

259 bulletin de l academie nationale de medecine.jn. 6829

260 medwave.jn. 814

261 diagnosis.jn. 514

262 bmj case reports.jn. 27404

263 bmj open quality.jn. 1036

264 einstein.jn. 1448

265 ceylon medical journal.jn. 2711

266 therapeutische umschau.jn. 7959

267 acta medica indonesiana.jn. 1056

268 annals of african medicine.jn. 871

269 dental & medical problems.jn. 321

270 new zealand medical journal.jn. 19142

271 clinical medicine & research.jn. 552

272 south african family practice.jn. 242

273 (journal of osteopathic medicine or journal of the american osteopathic association).jn. 8914

274 american journal of case reports.jn. 3326

275 wiener medizinische wochenschrift.jn. 13050

276 canadian journal of rural medicine.jn. 639

277 iranian journal of medical sciences.jn. 970

278 fukushima journal of medical science.jn. 775

279 nigerian postgraduate medical journal.jn. 1211

280 (romanian journal of internal medicine or medecine interne).jn. 1821

281 tidsskrift for den norske laegeforening.jn. 34097

282 international journal of health services.jn. 2334

283 agri dergisi.jn. 686

284 (journal of the royal college of physicians of edinburgh or proceedings of the royal college of physicians of edinburgh).jn. 1552

285 journal of investigative medicine high impact case reports.jn. 994

286 european journal of cardiovascular nursing.jn. 1506

287 neurology.jn. 43387

288 journal of medical ethics.jn. 5948

289 human molecular genetics.jn. 12361

290 translational behavioral medicine.jn. 1363

291 (journal of public health or journal of public health medicine).jn. 4228

292 laboratory medicine.jn. 905

293 health promotion international.jn. 2045

294 (occupational medicine oxford or transactions of the association of industrial medical officers or transactions of the society of occupational medicine or journal of the society of occupational medicine).jn. 4376

295 (european journal of preventive cardiology or journal of cardiovascular risk or “european journal of cardiovascular prevention & rehabilitation“).jn. 4891

296 american journal of clinical pathology.jn. 18282

297 cardiovascular research.jn. 11950

298 european journal of hospital pharmacy science & practice.jn. 914

299 bmj supportive & palliative care.jn. 1688

300 european heart journal acute cardiovascular care.jn. 1133

301 american journal of hypertension.jn. 7847

302 medical humanities.jn. 980

303 bmj leader.jn. 57

304 british journal of sports medicine.jn. 7432

305 bmj open ophthalmology.jn. 415

306 jnci cancer spectrum.jn. 599

307 international journal of health policy & management.jn. 1568

308 international journal of epidemiology.jn. 9306

309 (“bmj quality & safety” or “quality in health care” or “quality & safety in health care“).jn. 3622

310 bosnian journal of basic medical sciences.jn. 1273

311 thorax.jn. 13877

312 british journal of ophthalmology.jn. 21522

313 (allergy or acta allergologica).jn. 11256

314 alcohol & alcoholism.jn. 3625

315 stroke & vascular neurology.jn. 396

316 europace.jn. 6605

317 european heart journal cardiovascular pharmacotherapy.jn. 612

318 european heart journal quality of care & clinical outcomes.jn. 574

319 lancet planetary health.jn. 893

320 journal of pharmacy & pharmacology.jn. 15387

321 european heart journal cardiovascular imaging.jn. 3282

322 archivos argentinos de pediatria.jn. 2923

323 annals of behavioral medicine.jn. 2061

324 journals of gerontology series a biological sciences & medical sciences.jn. 6047

325 journal of the national cancer institute.jn. 22867

326 journal of medical genetics.jn. 8946

327 journal of nutrition.jn. 22145

328 (sexually transmitted infections or genitourinary medicine or british journal of venereal diseases).jn. 8837

329 bmj paediatrics open.jn. 757

330 schizophrenia bulletin.jn. 4709

331 inflammatory bowel diseases.jn. 6759

332 health promotion journal of australia.jn. 1213

333 european journal of public health.jn. 3909

334 advances in nutrition.jn. 1427

335 age & ageing.jn. 6249

336 journal of crohns & colitis.jn. 2837

337 international journal of gynecological cancer.jn. 6284

338 ebiomedicine.jn. 4423

339 (cin computers informatics nursing or computers in nursing).jn. 2346

340 mbio.jn. 6220

341 family medicine & community health.jn. 185

342 gerontologist.jn. 6196

343 acta orthopaedica et traumatologica turcica.jn. 1928

344 (“bmj sexual & reproductive health” or british journal of family planning or “journal of family planning & reproductive health care“).jn. 2128

345 (“journal of epidemiology & community health” or “british journal of social medicine” or “british journal of preventive & social medicine“).jn. 7815

346 gaceta sanitaria.jn. 3461

347 nephrology dialysis transplantation.jn. 18729

348 veterinary record.jn. 36161

349 turk kardiyoloji dernegi arsivi.jn. 2400

350 (“occupational & environmental medicine” or “british journal of industrial medicine“).jn. 7772

351 (rheumatology or annals of physical medicine or “rheumatology & physical medicine” or “rheumatology & rehabilitation” or “british journal of rheumatology“).jn. 15875

352 journal of neurology neurosurgery & psychiatry.jn. 18218

353 nephrologie et therapeutique.jn. 1352

354 annals of the royal college of surgeons of england.jn. 12167

355 physical therapy.jn. 7403

356 journal of medical imaging & radiation sciences.jn. 1136

357 brain.jn. 11338

358 british journal of clinical pharmacology.jn. 13061

359 nursing inquiry.jn. 1194

360 european journal of cardio thoracic surgery.jn. 14425

361 tobacco control.jn. 4109

362 (heart or british heart journal).jn. 20552

363 “jmir public health and surveillance“.jn. 1083

364 western journal of emergency medicine.jn. 2416

365 global heart.jn. 725

366 cadernos de saude publica.jn. 6979

367 bulletin of the world health organization.jn. 11239

368 gastroenterology nursing.jn. 2017

369 (annals of global health or journal of the mount sinai hospital new york or mount sinai journal of medicine).jn. 6114

370 revista de saude publica.jn. 5039

371 asia pacific journal of public health.jn. 2561

372 (“bjog an international journal of obstetrics & gynaecology” or “journal of obstetrics & gynaecology of the british commonwealth” or “journal of obstetrics & gynaecology of the british empire” or “british journal of obstetrics & gynaecology“).jn. 19208

373 british dental journal.jn. 20185

374 international journal of nursing studies.jn. 4710

375 journal of child health care.jn. 1016

376 international journal of older people nursing.jn. 826

377 international journal of gynaecology & obstetrics.jn. 11788

378 international nursing review.jn. 2837

379 sleep.jn. 7010

380 nicotine & tobacco research.jn. 4749

381 (journal of tropical pediatrics or “journal of tropical pediatrics & african child health” or “journal of tropical pediatrics & environmental child health“).jn. 4895

382 physical therapy.jn. 7403

383 nutrition reviews.jn. 8575

384 health policy & planning.jn. 2519

385 human reproduction.jn. 15742

386 (medical mycology or sabouraudia or “journal of medical & veterinary mycology“).jn. 4751

387 journal of the american medical informatics association.jn. 4161

388 international journal of pharmacy practice.jn. 1026

389 american journal of clinical nutrition.jn. 21852

390 (american journal of health system pharmacy or american journal of hospital pharmacy).jn. 17478

391 rmd open.jn. 1083

392 evidence based nursing.jn. 2176

393 journal of clinical pathology.jn. 16609

394 (emergency medicine journal or “journal of accident & emergency medicine” or archives of emergency medicine).jn. 7878

395 evidence based mental health.jn. 1791

396 (“bmj health & care informatics” or “informatics in primary care” or “journal of innovation in health informatics“).jn. 749

397 bmj open gastroenterology.jn. 476

398 bmj open respiratory research.jn. 693

399 annals of the rheumatic diseases.jn. 16714

400 bmj global health.jn. 2591

401 archives of disease in childhood.jn. 21159

402 injury prevention.jn. 2699

403 indian journal of medical ethics.jn. 1304

404 (“journal of health population & nutrition” or journal of diarrhoeal diseases research).jn. 1825

405 glob health action.jn. 1885

406 or/75-405 3025414

407 73 and 406 3691

408 74 and 406 5206

409 limit 407 to yr=“1860 - 2022” 3567

410 limit 408 to yr=“1860 - 2022” 5201

https://ovidsp.ovid.com/ovidweb.cgi?T=JS&NEWS=N&PAGE=main&SHAREDSEARCHID=5aG3iSbGHuDgw66zcflKjOPKK2JgfXFH68hUptypX9Je47onfxPKkhsuGtXZNXAWi

#### Database

Ovid MEDLINE(R) ALL <1946 to January 25, 2023>

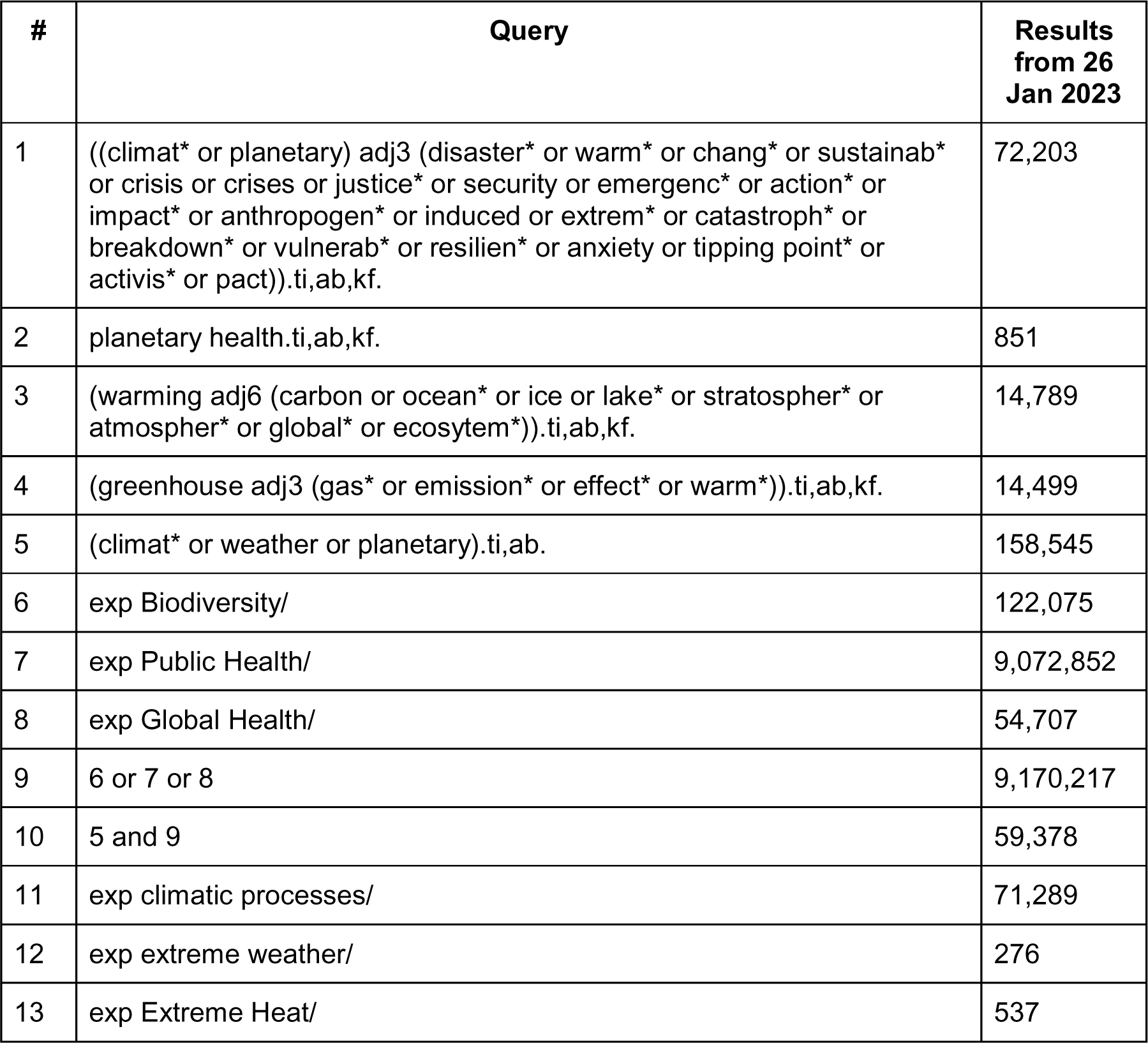
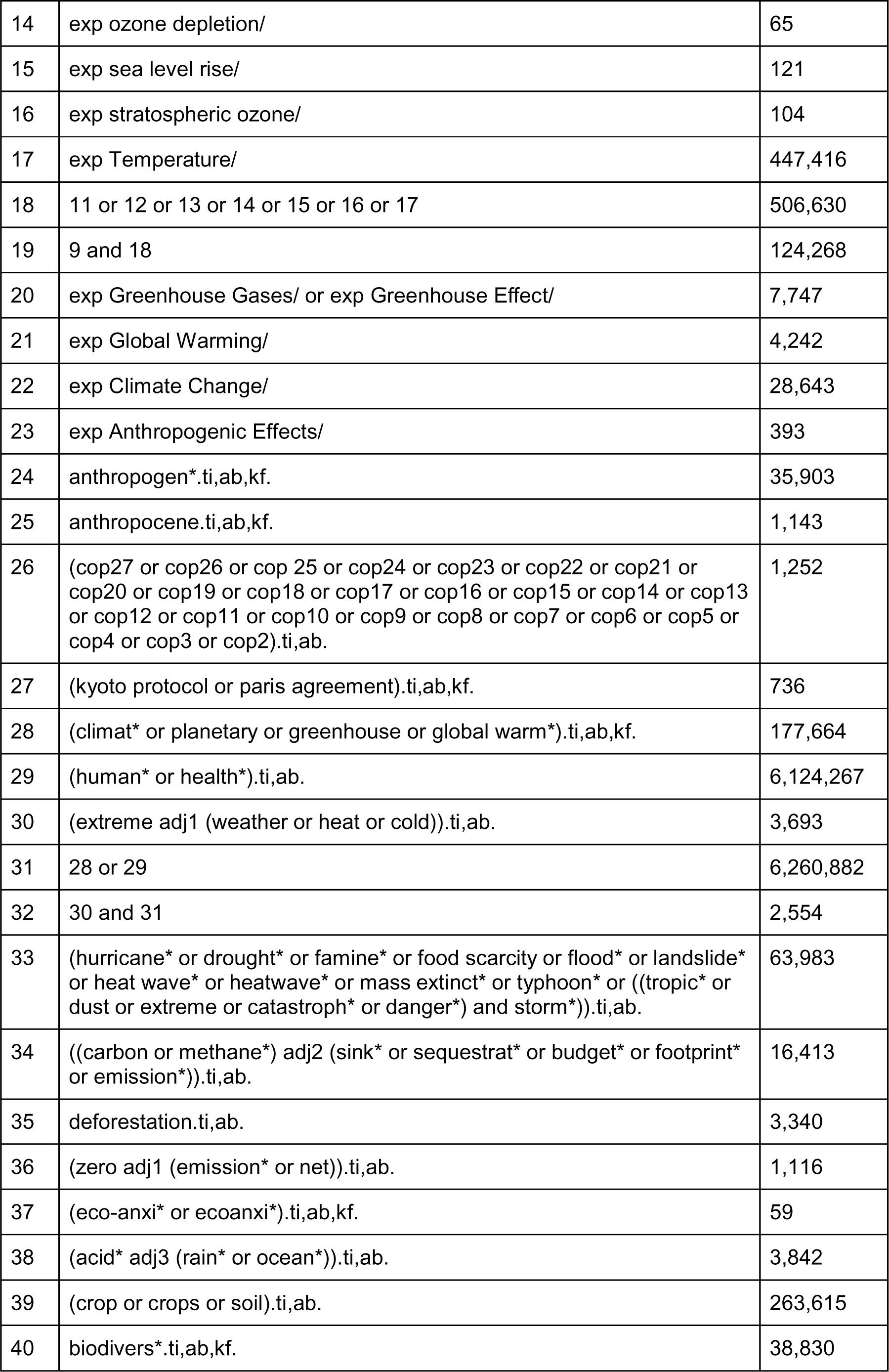
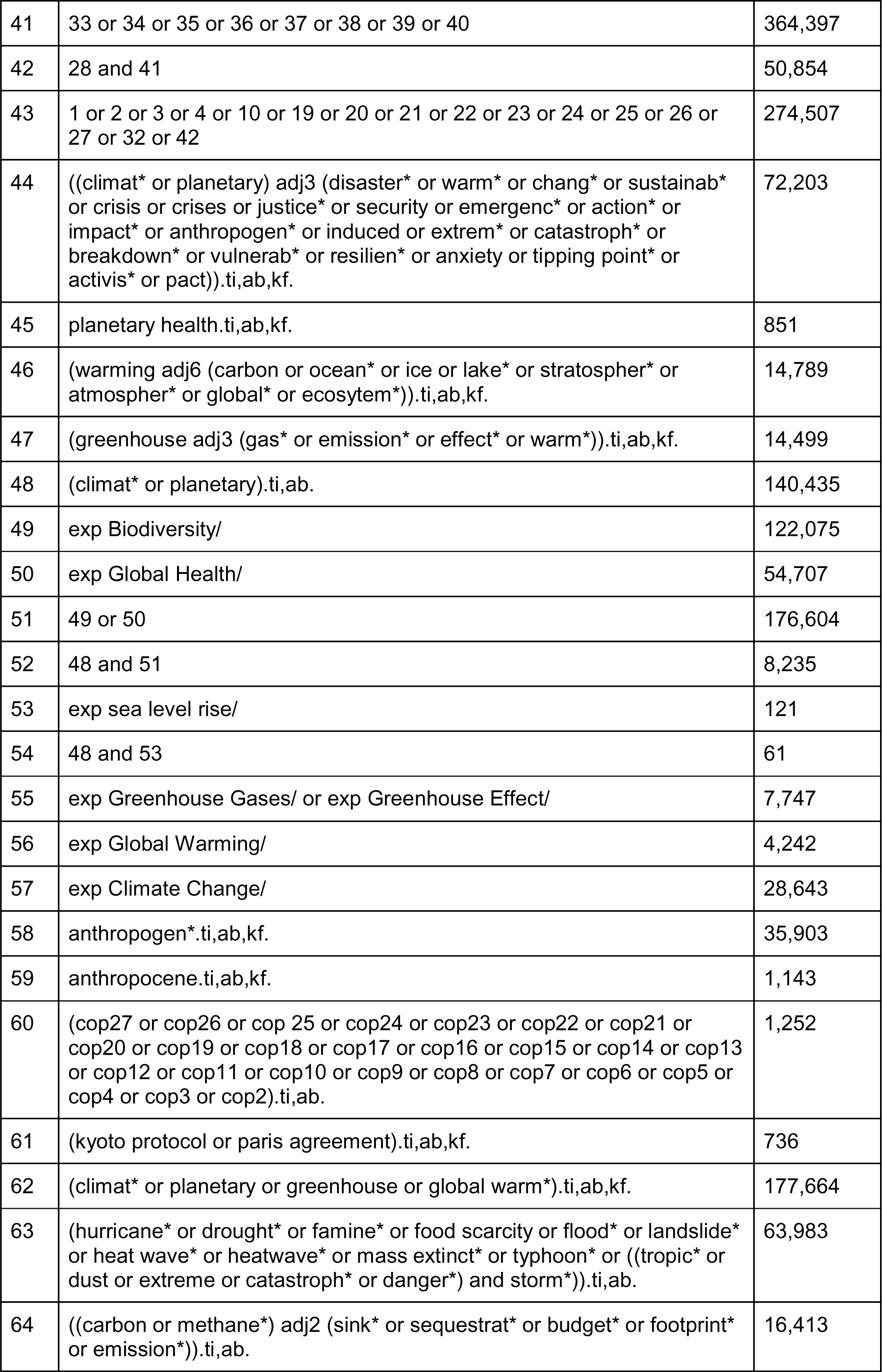
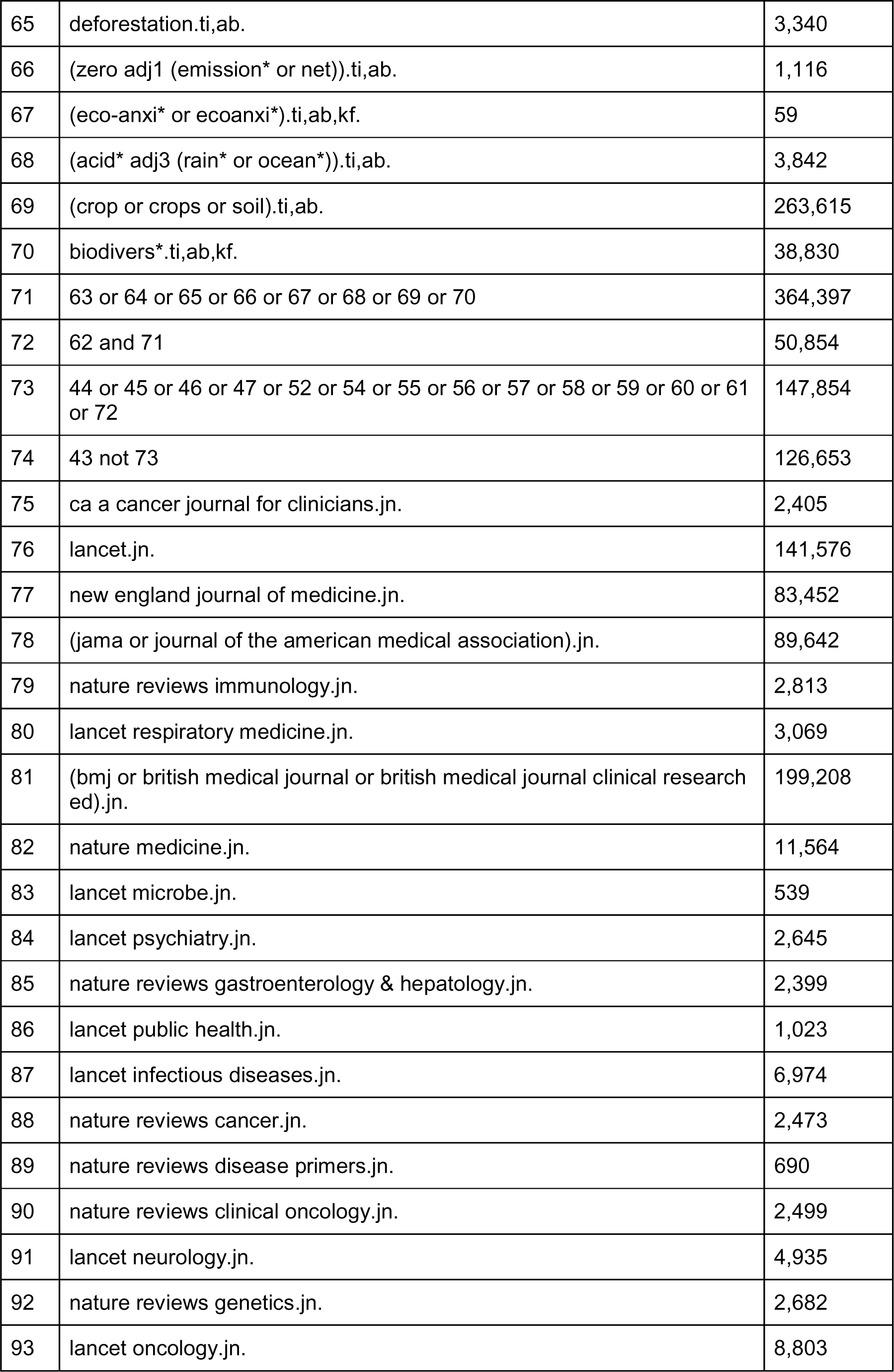
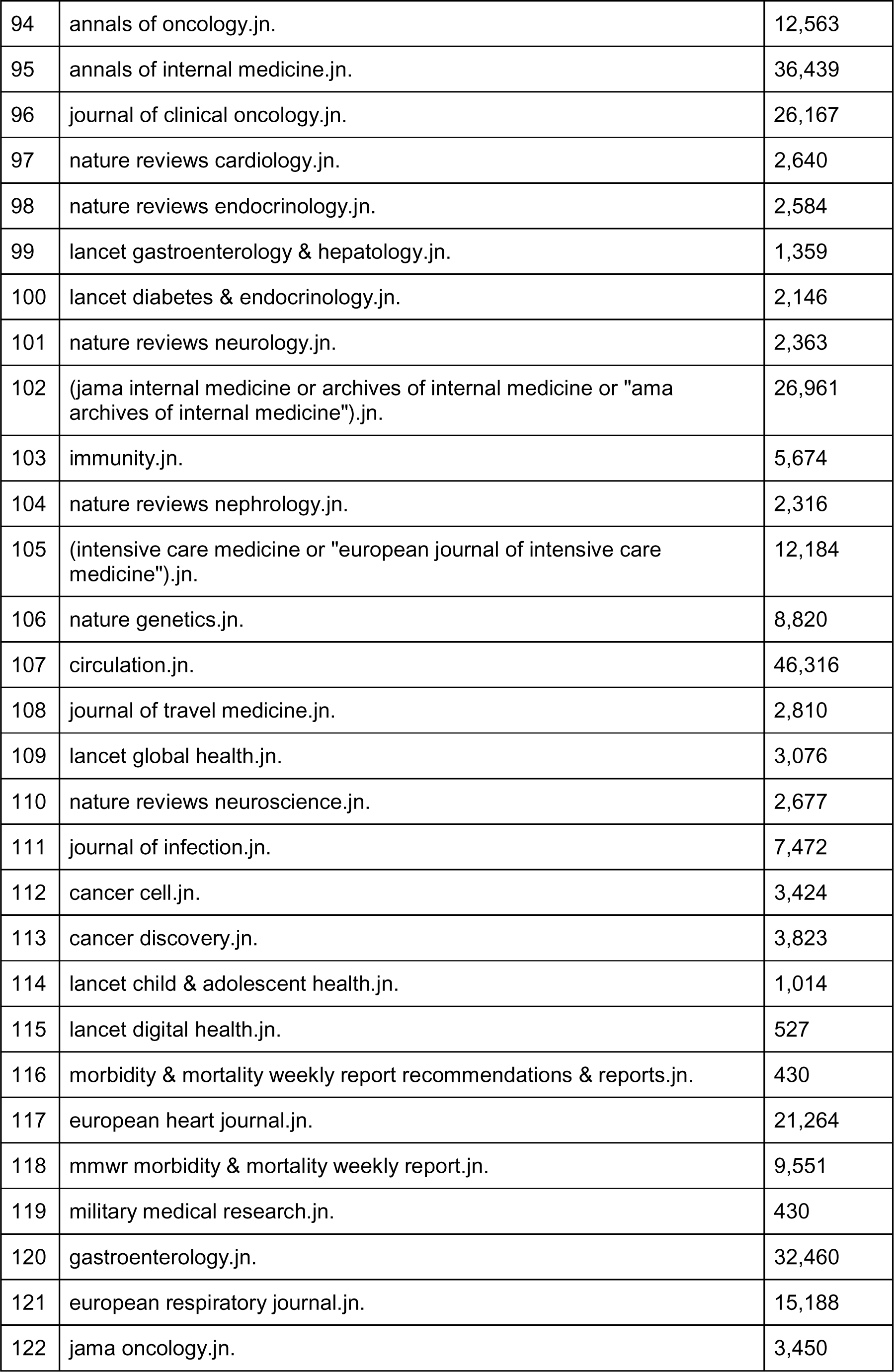
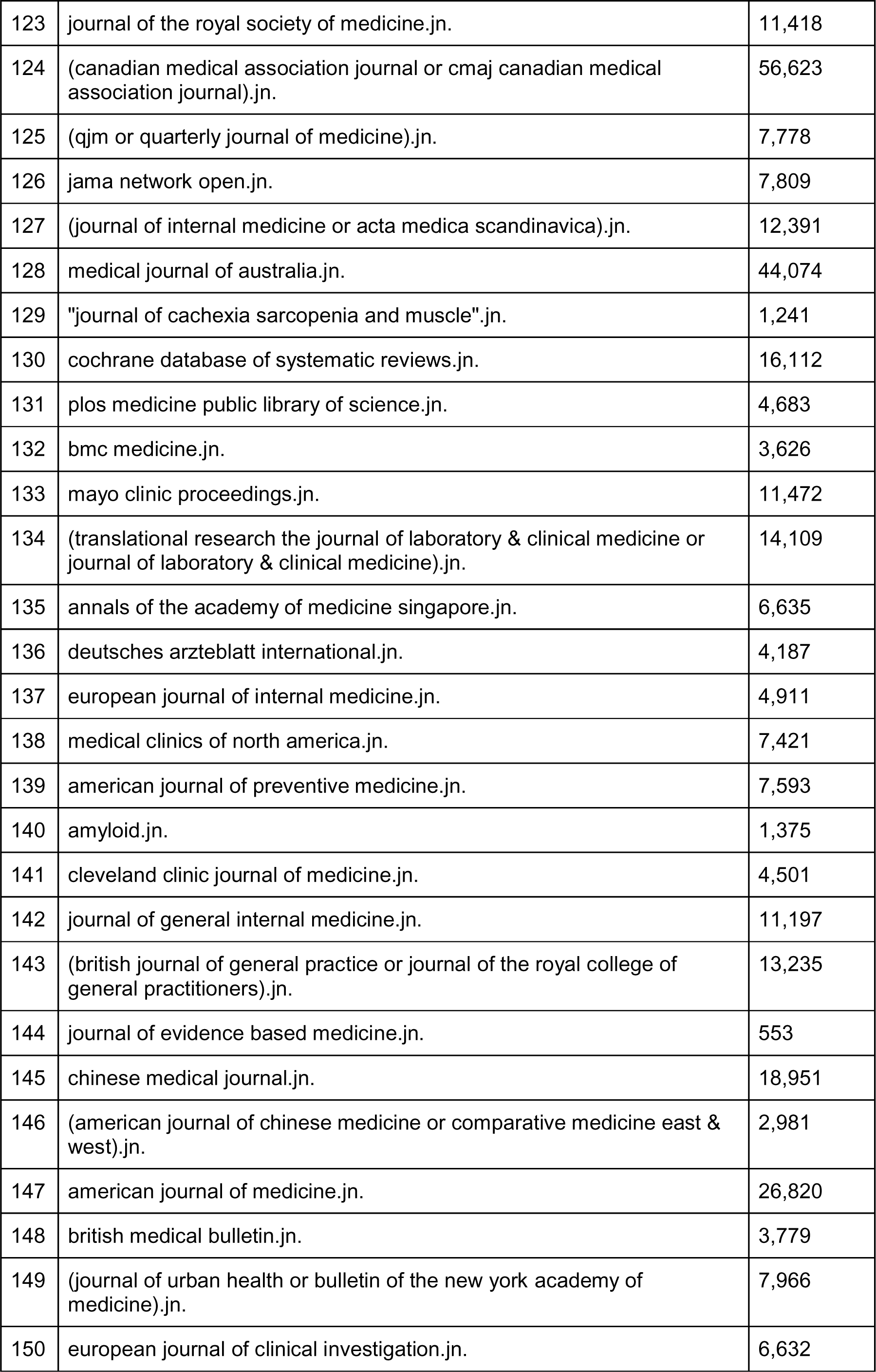
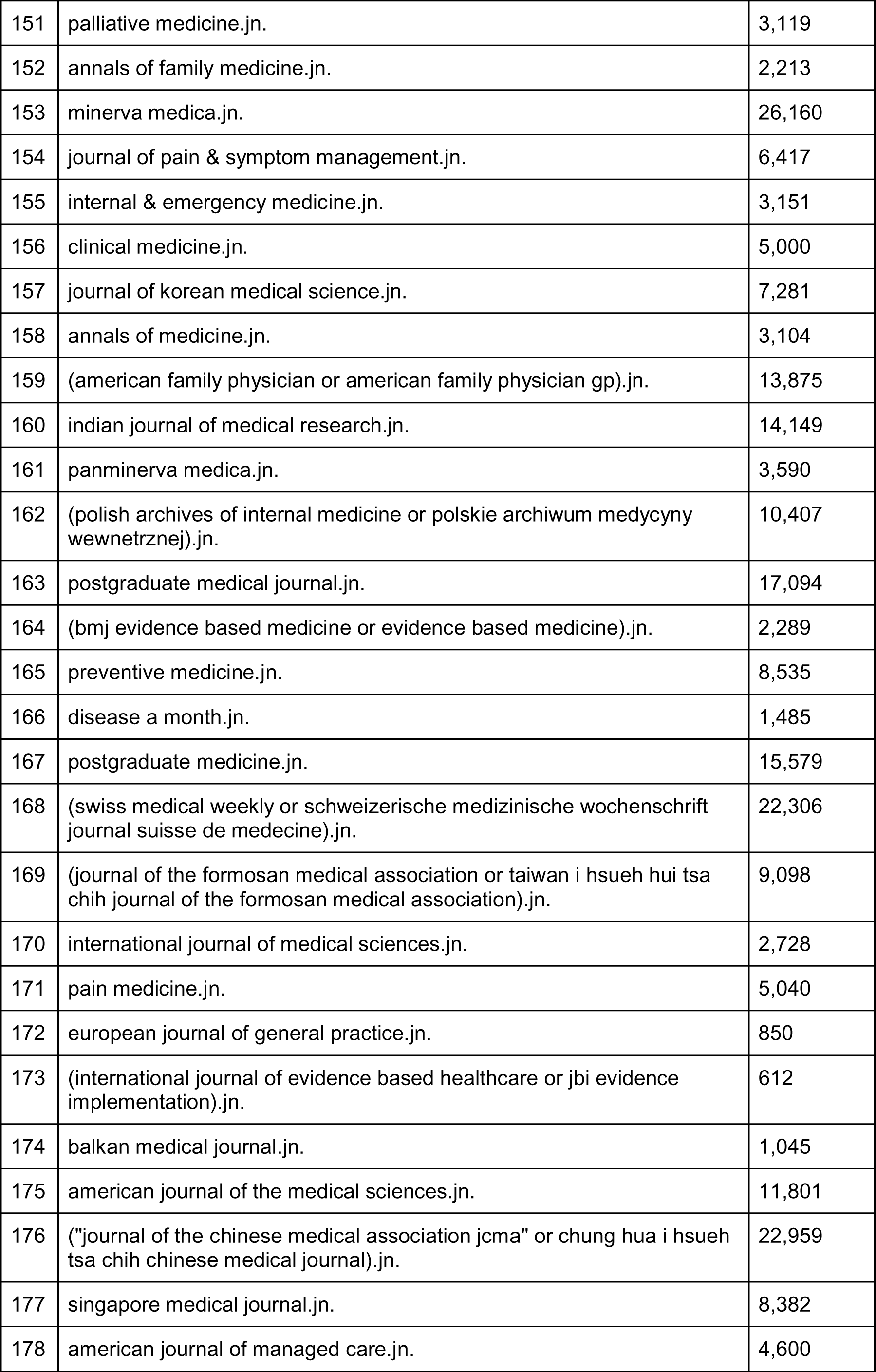
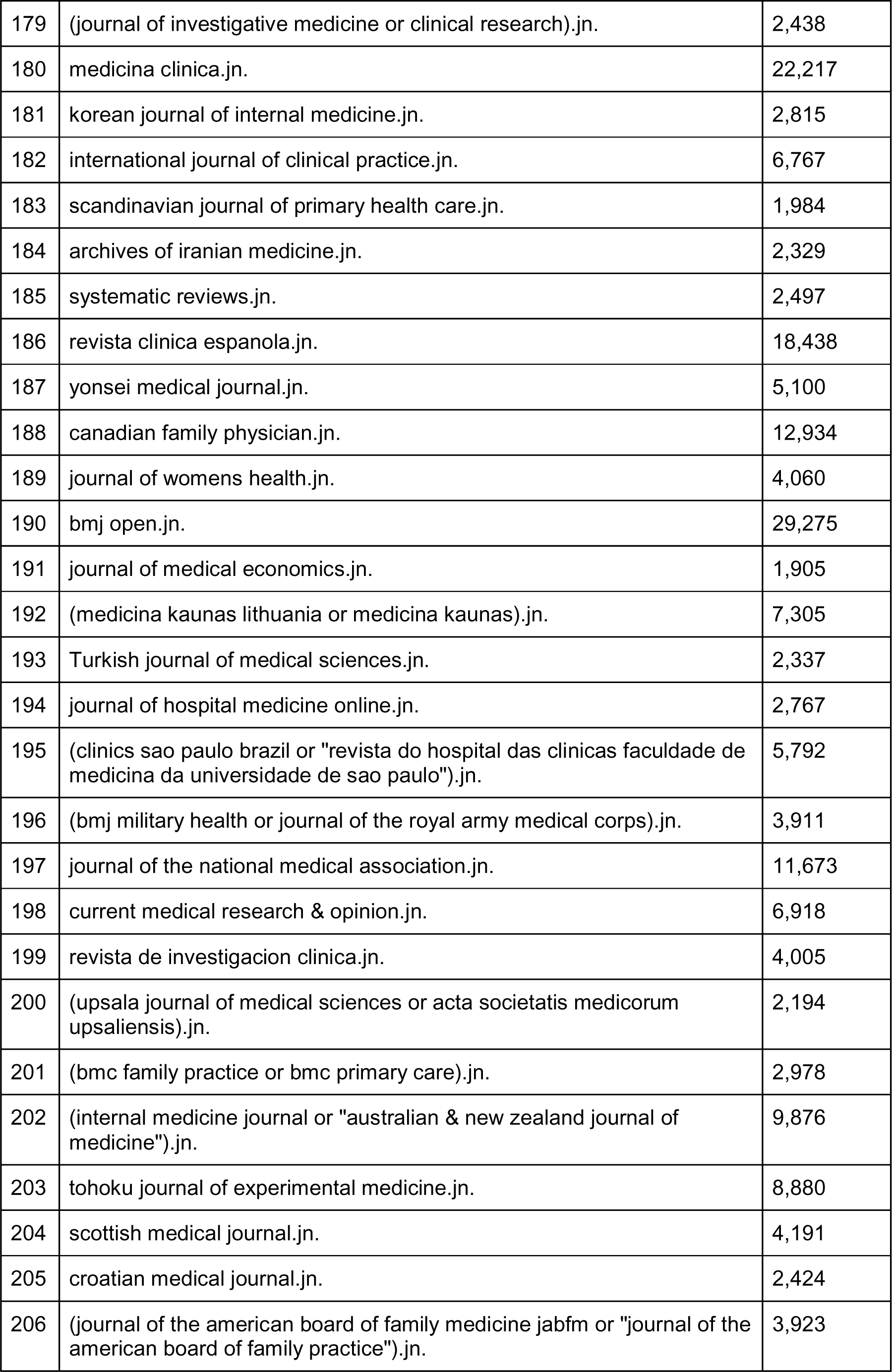
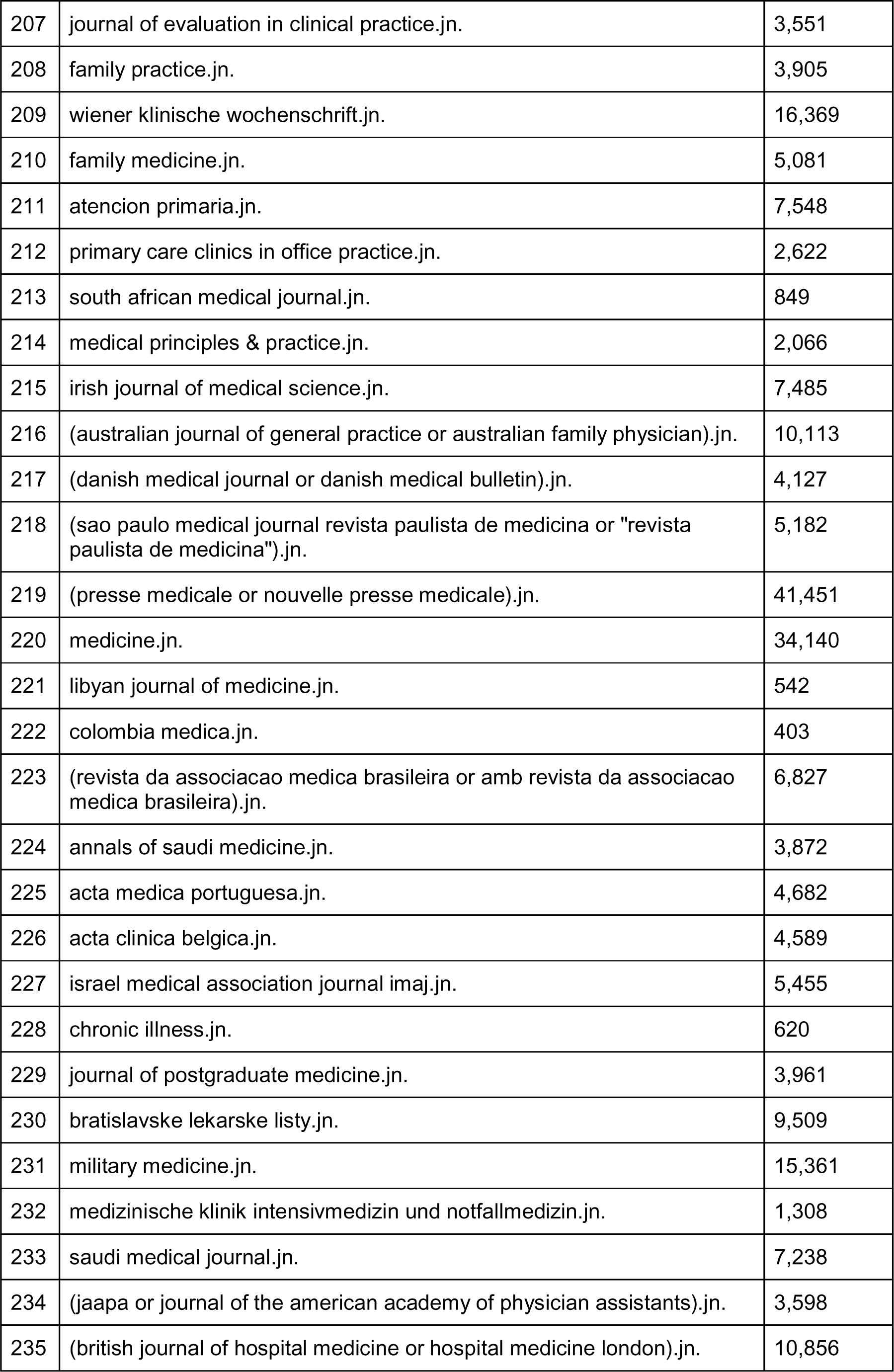
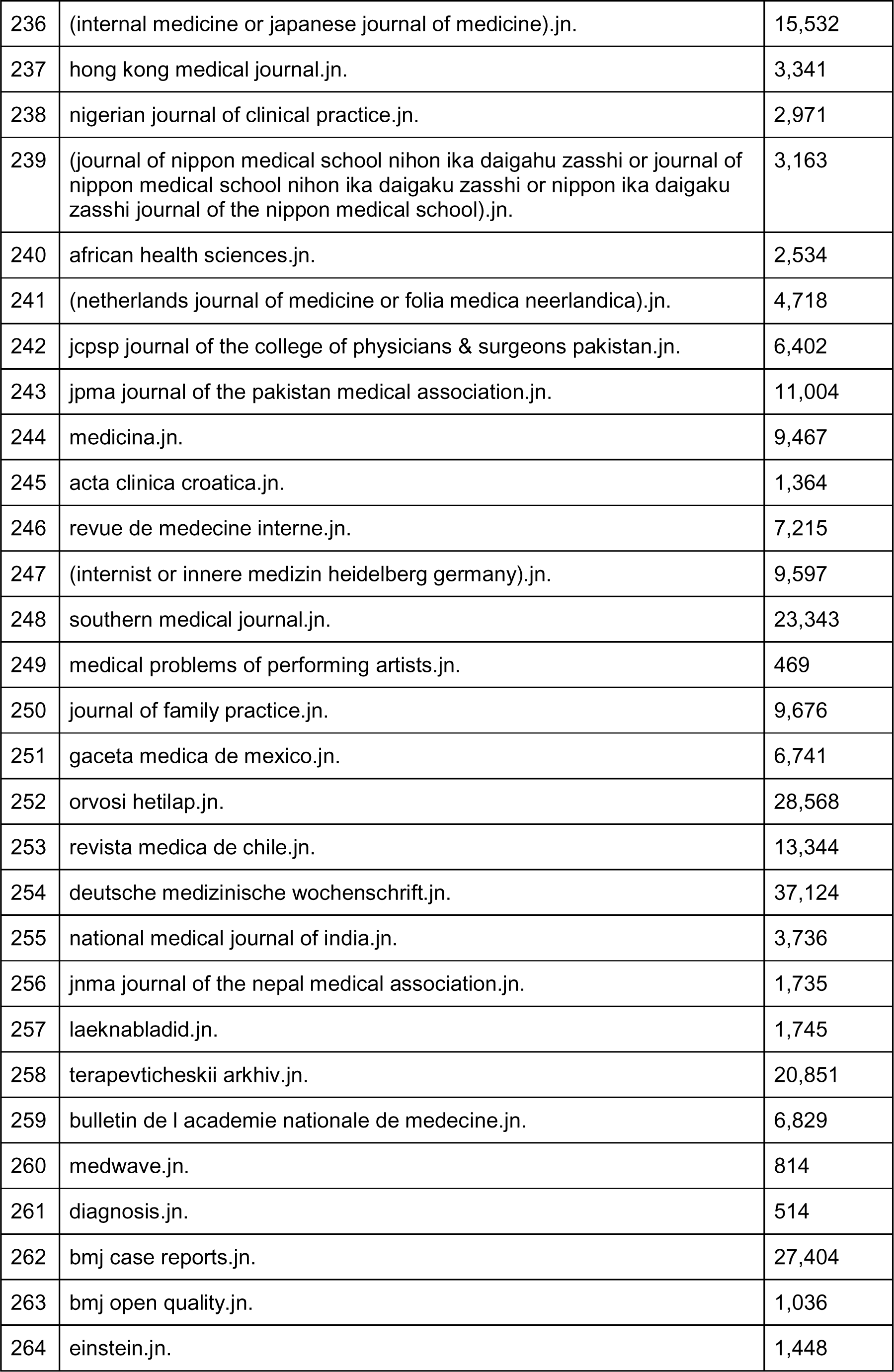
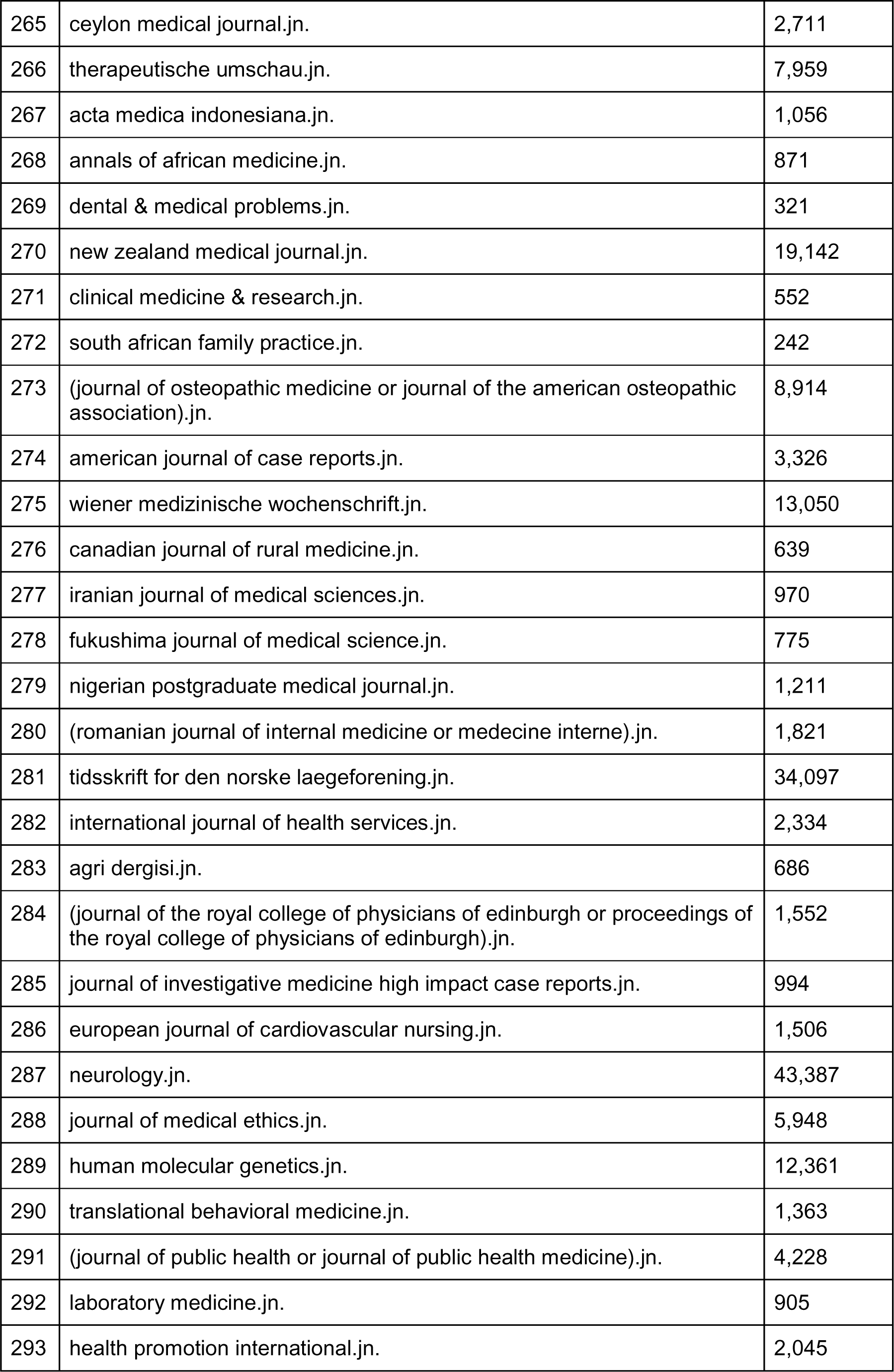
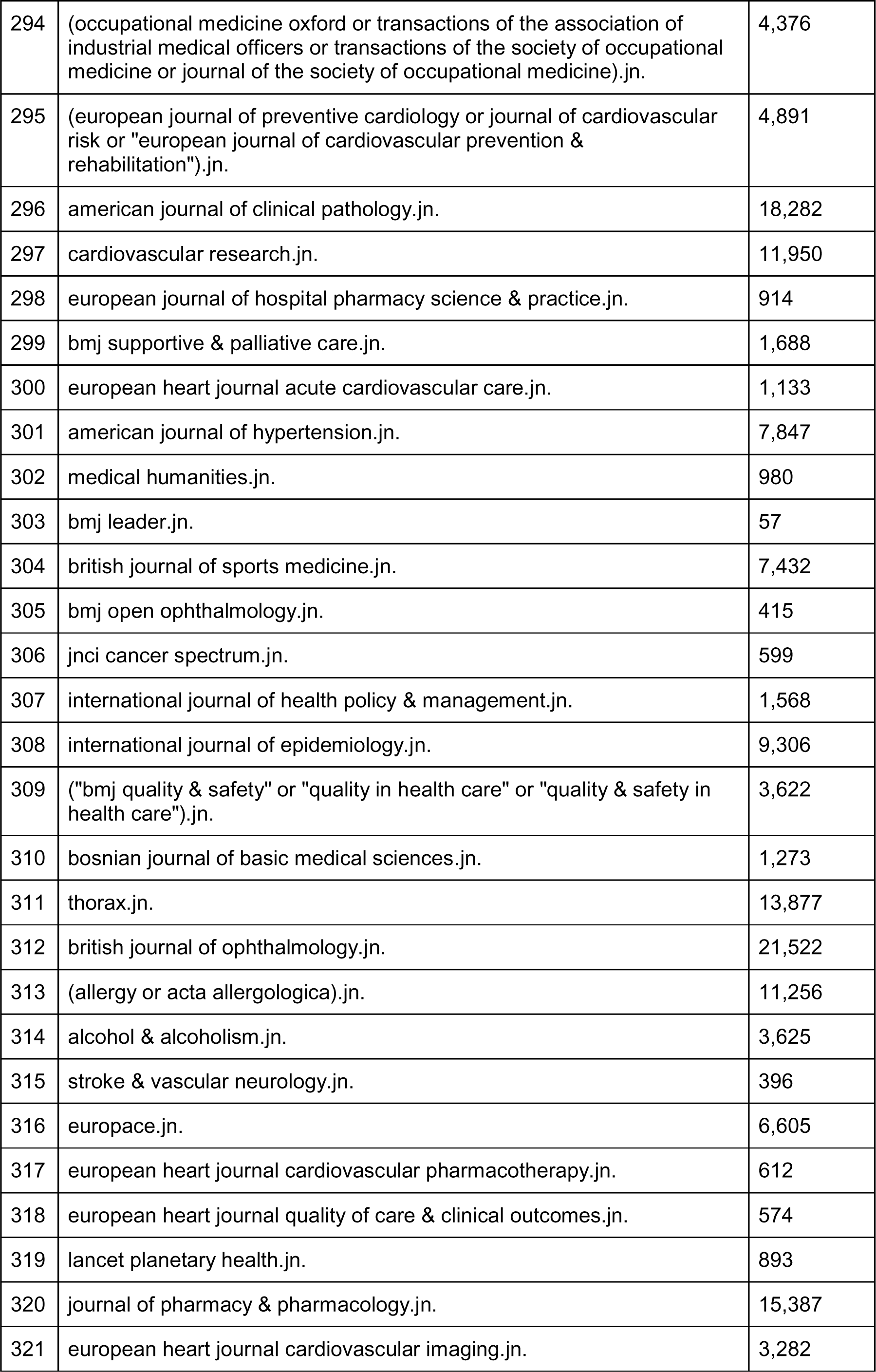
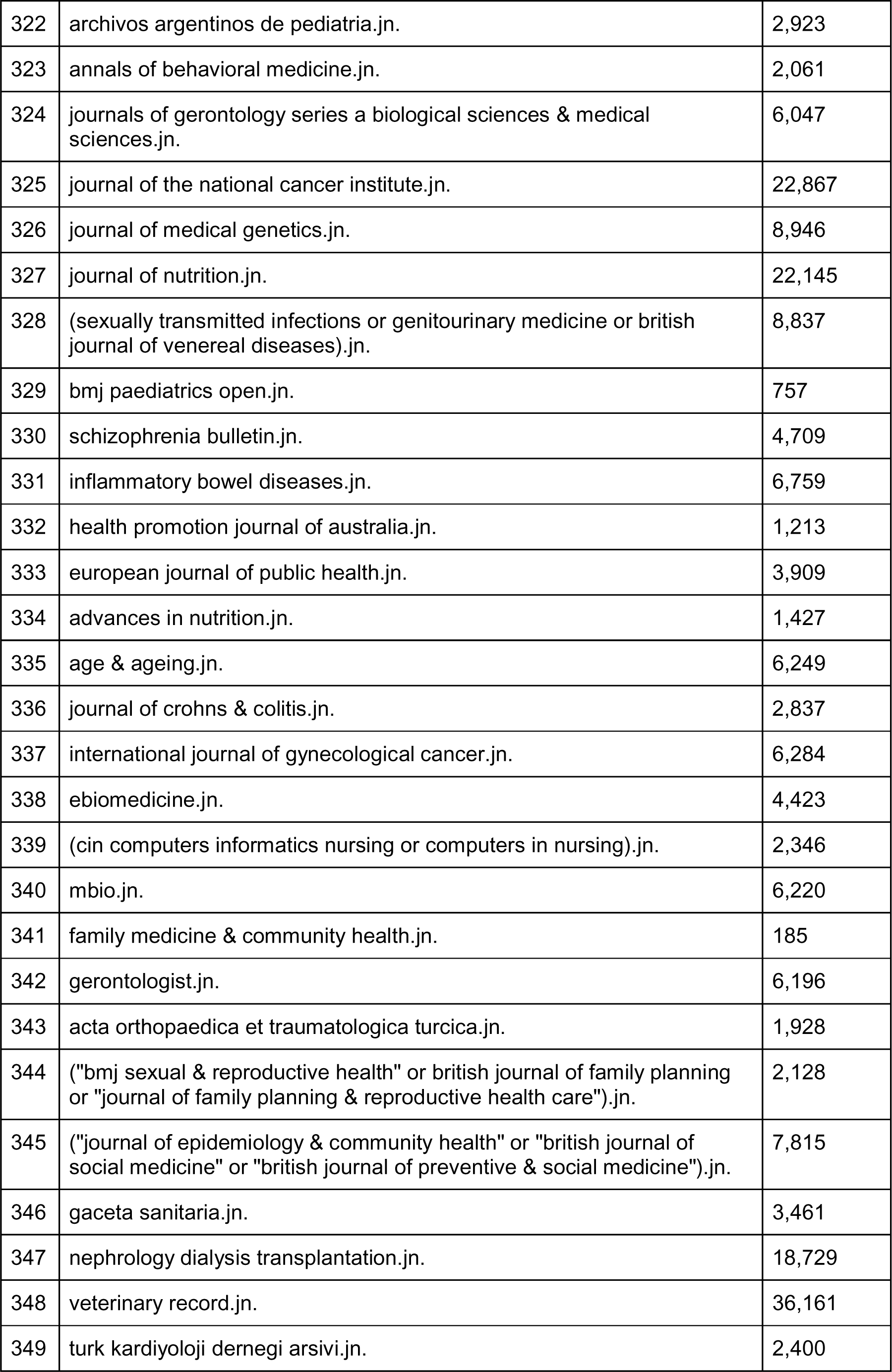
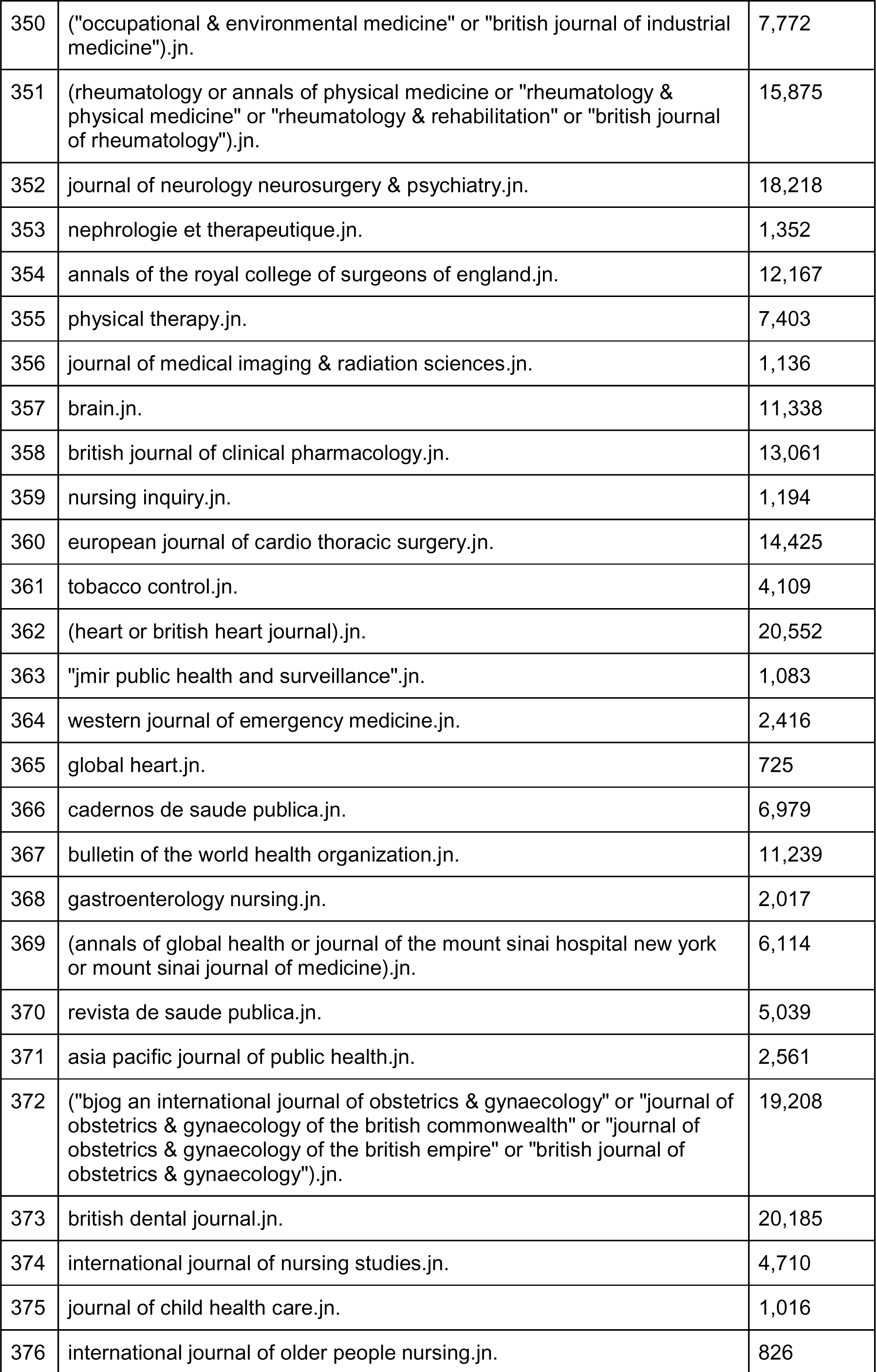
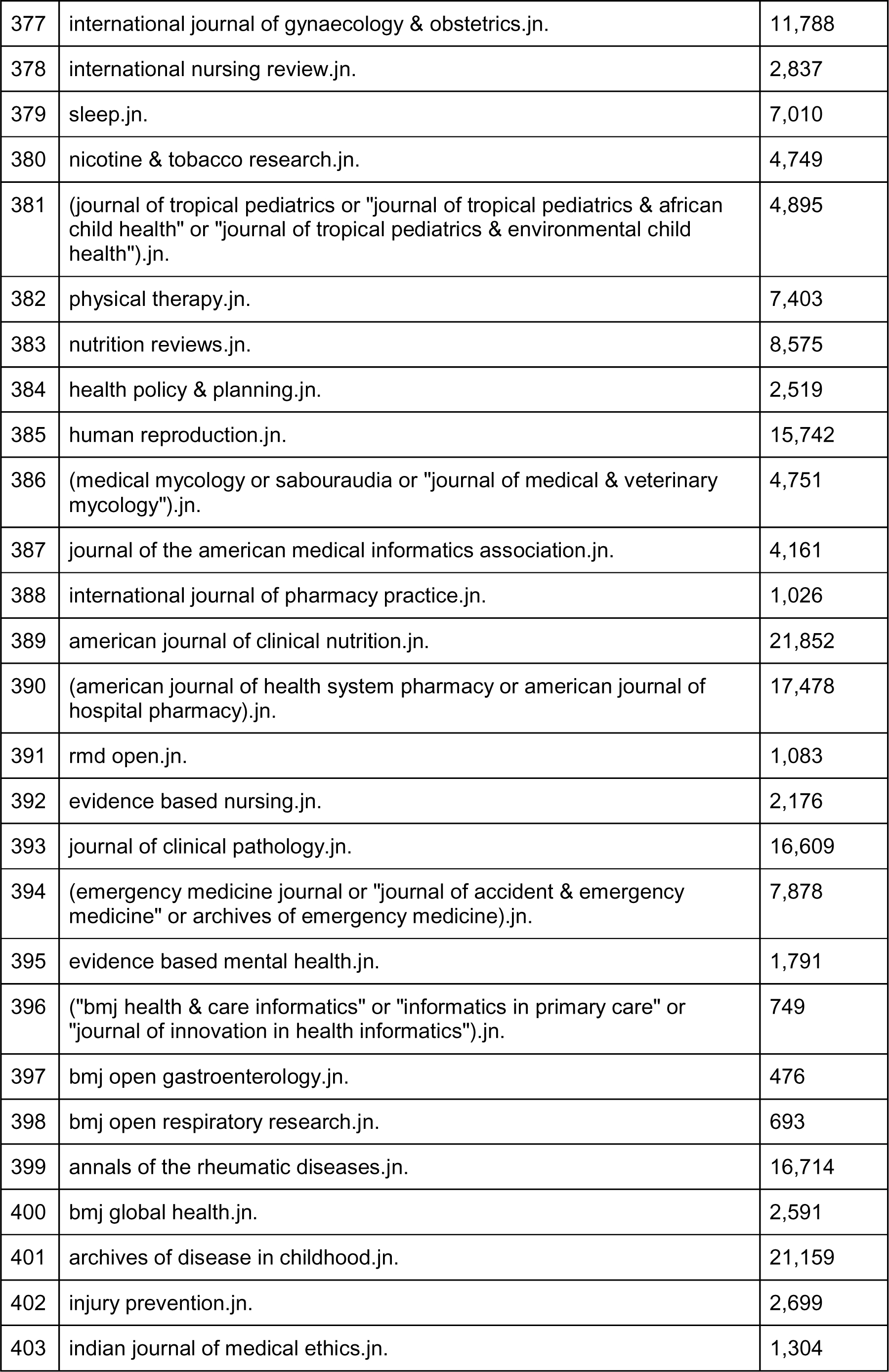
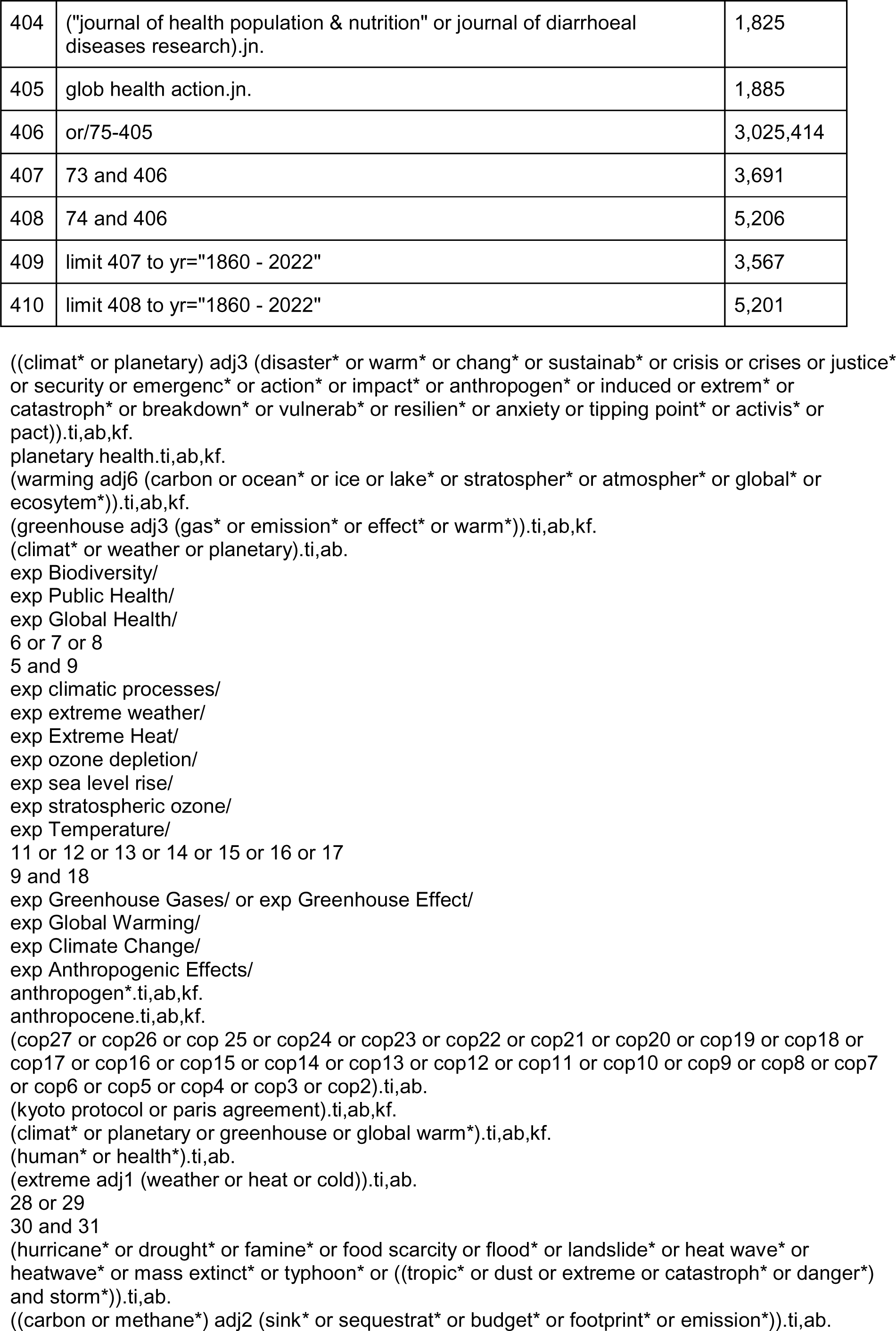
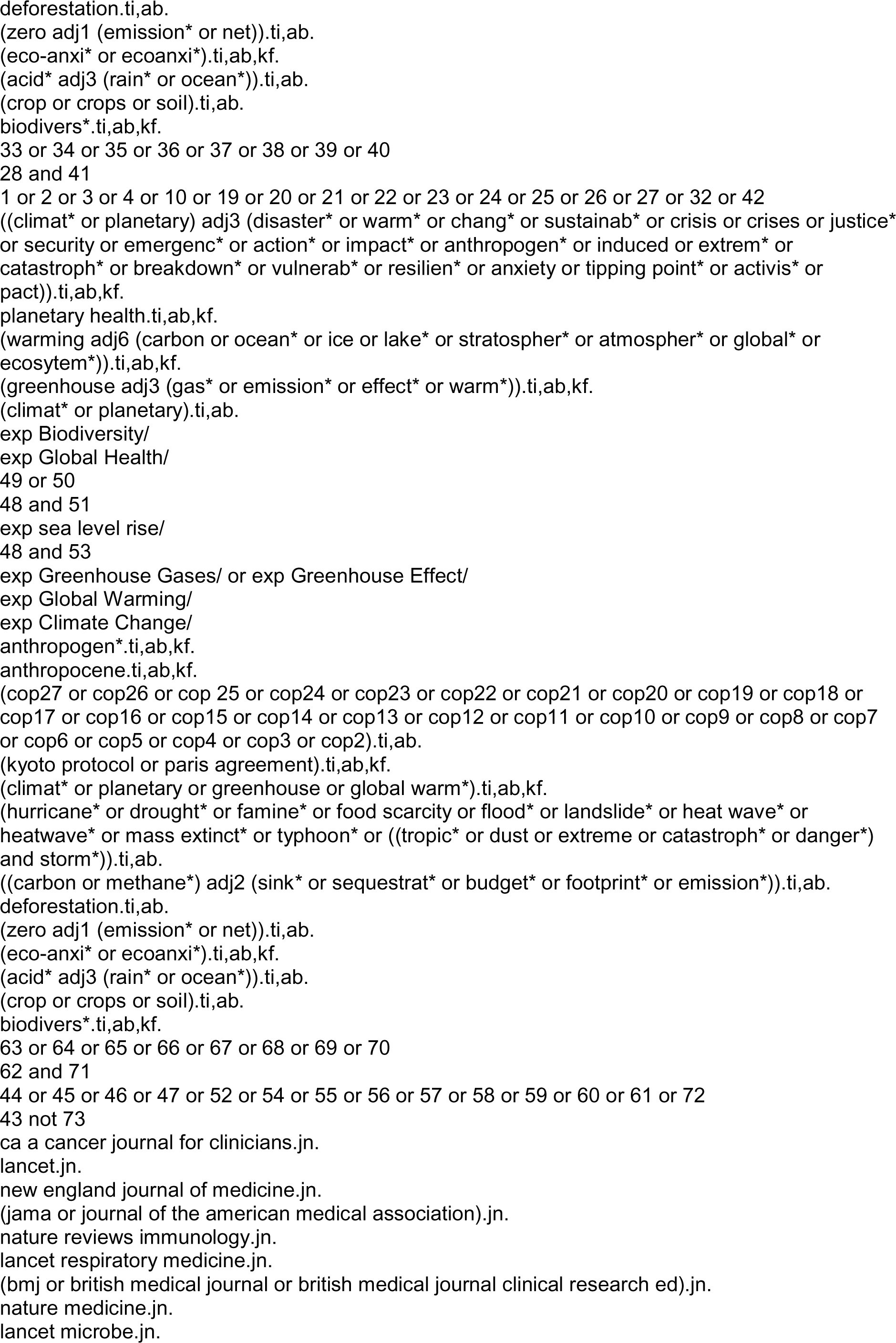
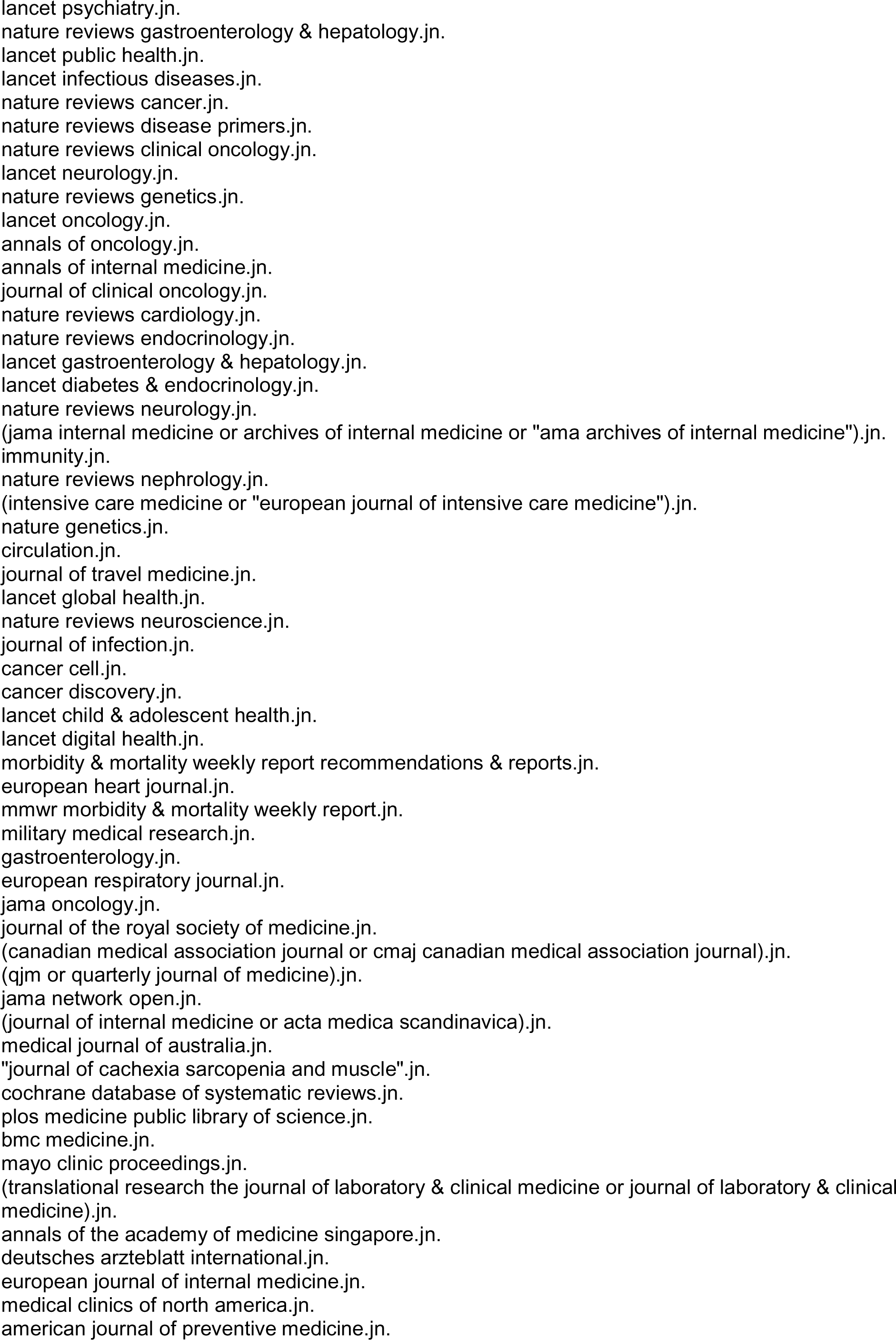
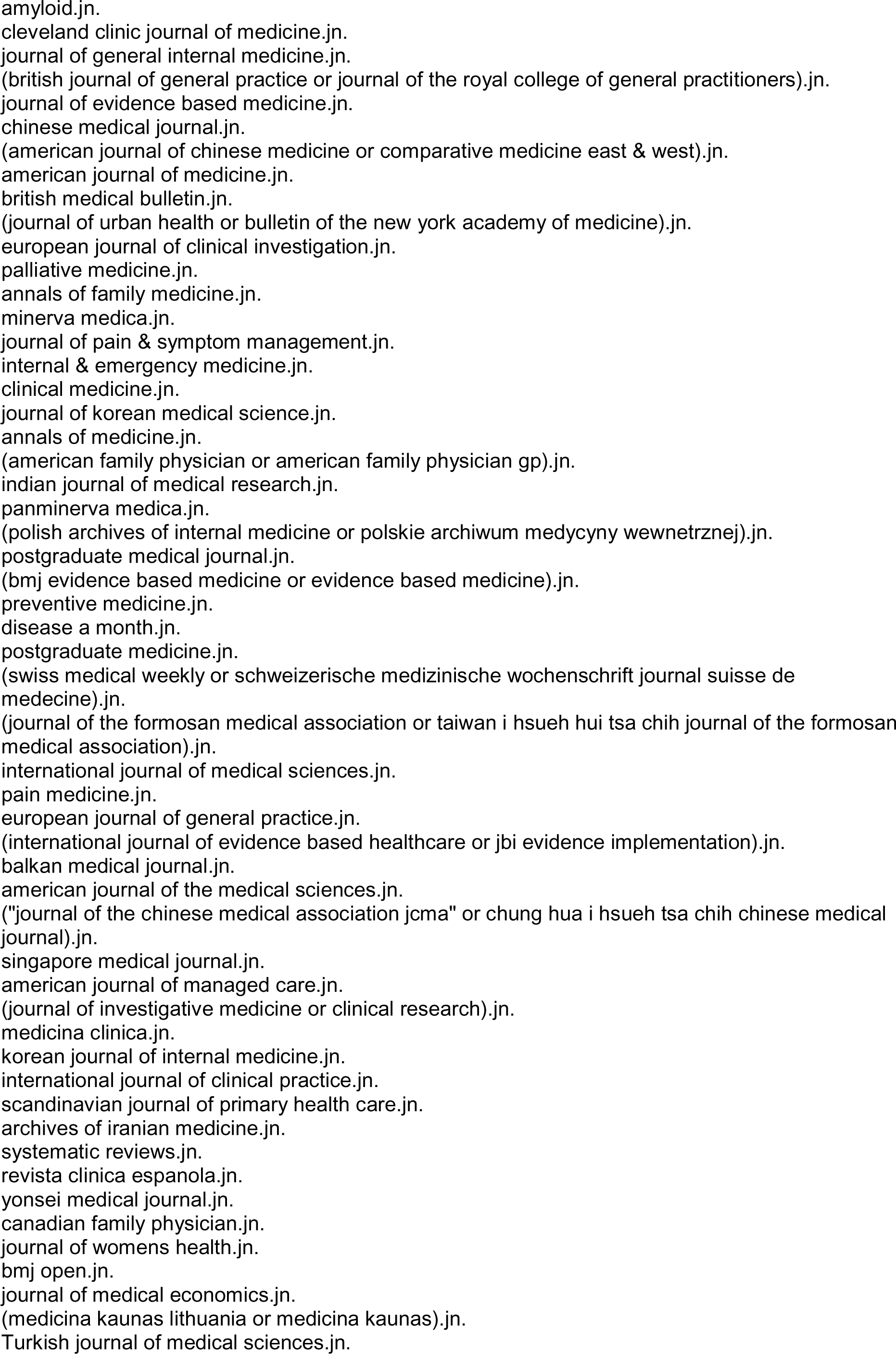
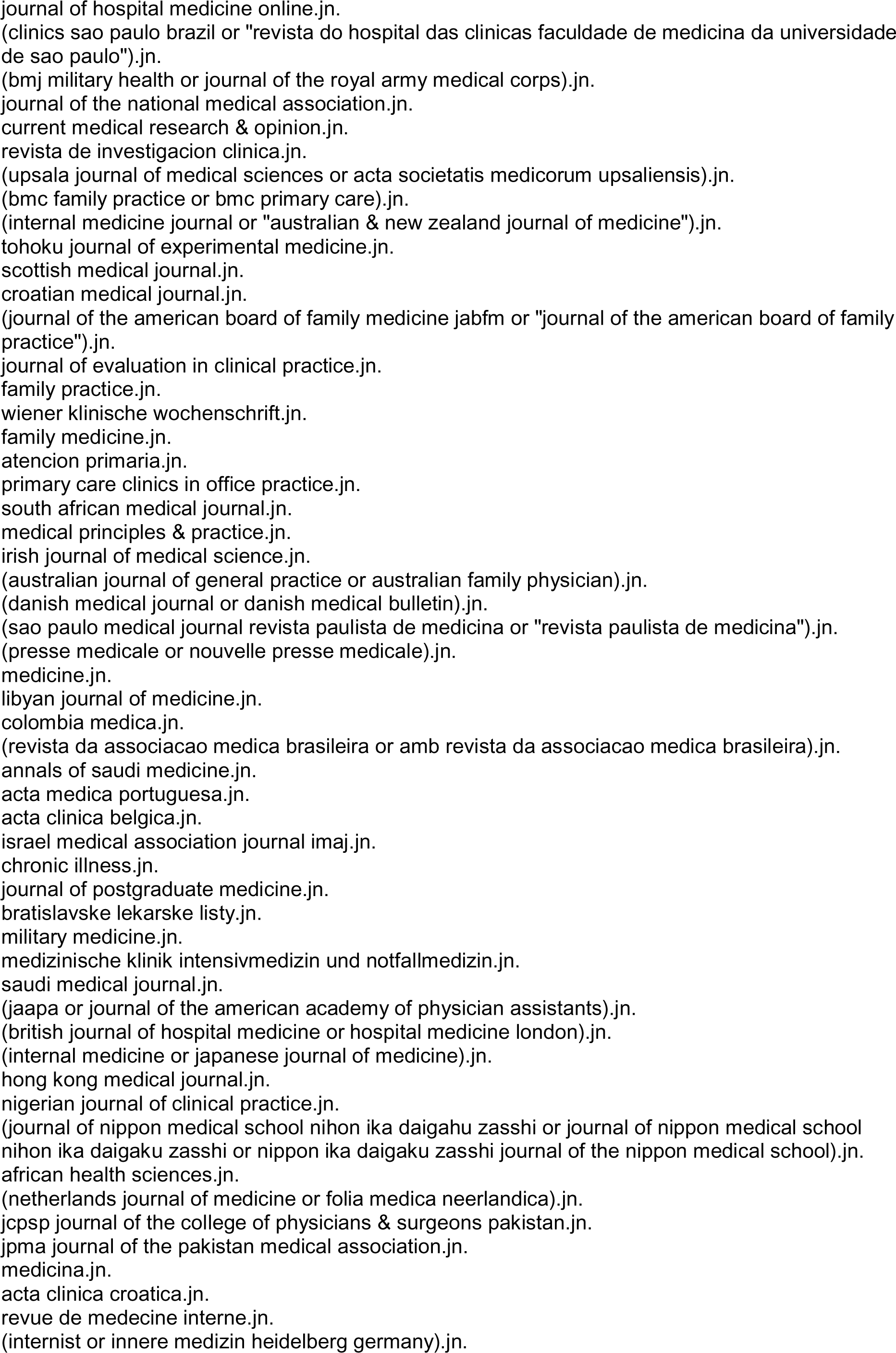
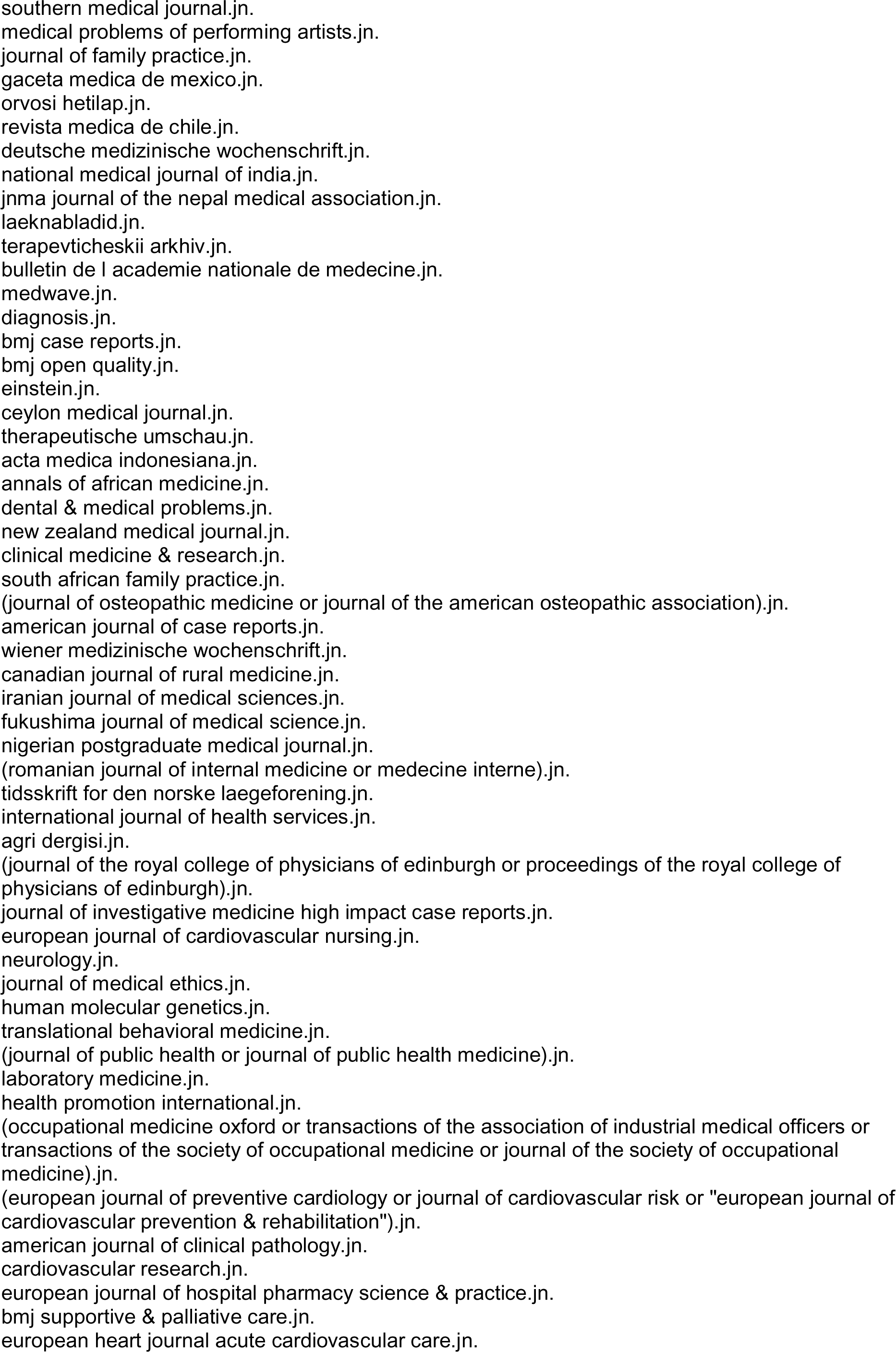
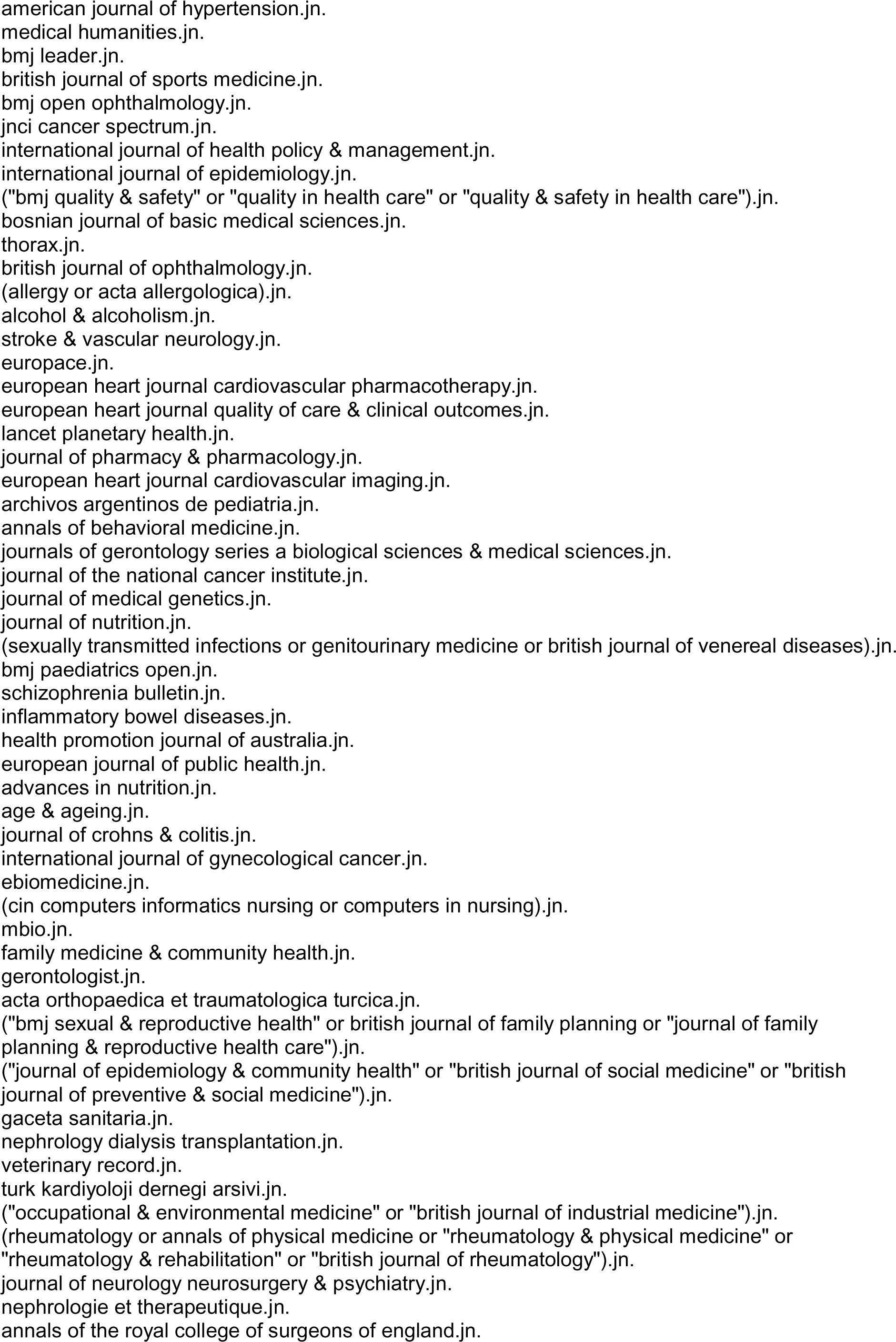
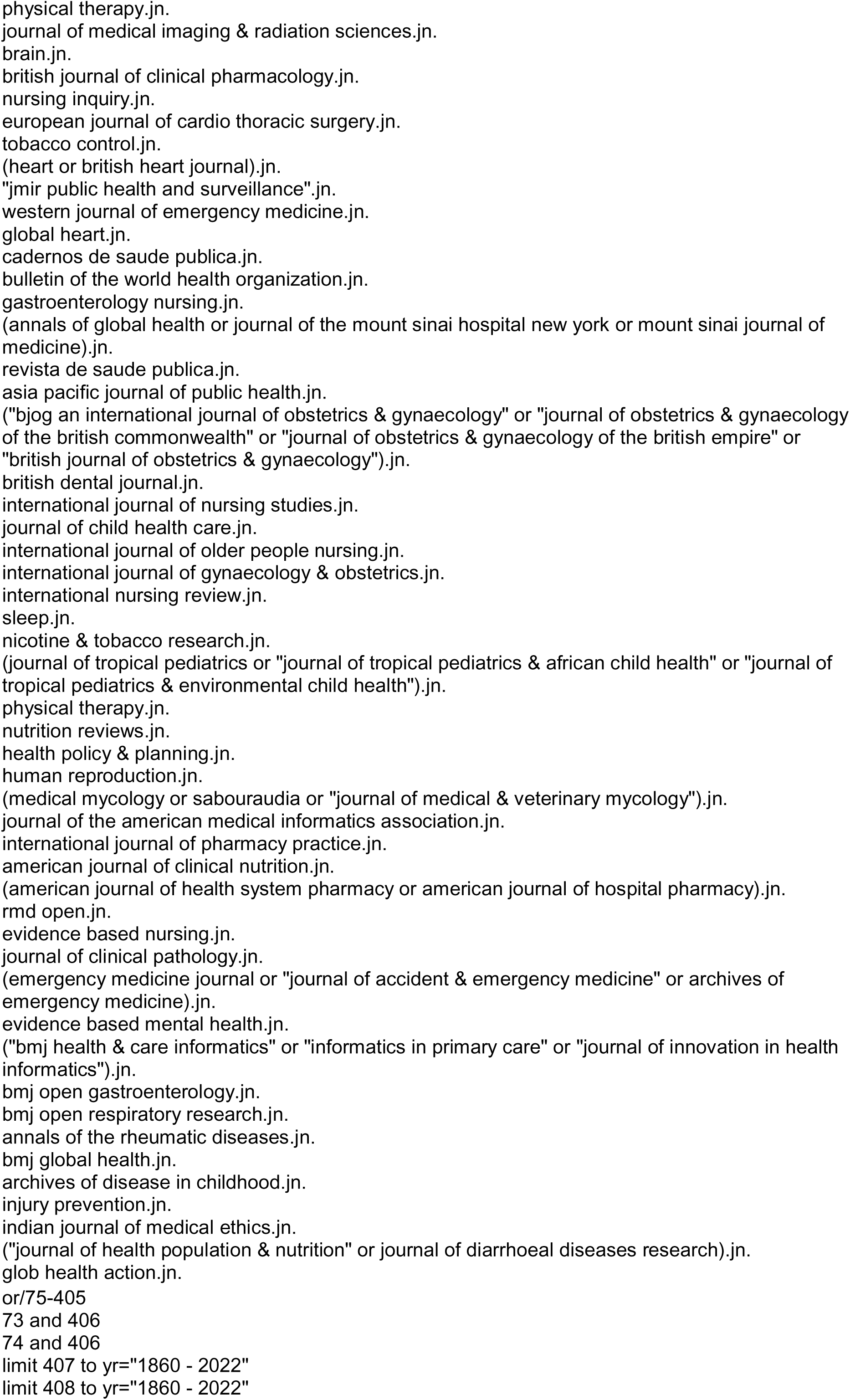

https://ovidsp.ovid.com/ovidweb.cgi?T=JS&NEWS=N&PAGE=main&SHAREDSEARCHID=36B5EVfYAAfErqmPSUVB4zXyLraTGkTg1cBDQKFX41YQZ29cpCArfA5xn2ZvRhRGm

### Appendix 2 Coverage of climate change and human health: Article inclusion and exclusion criteria to apply to articles found in the original MEDLINE search

#### Process

Start by screening the title and make a decision according to the criteria below. If you can’t decide based on the title, go to the abstract to make a decision. Do NOT go to full text even if it is available.

#### Inclusion criteria

Include all articles that mention the following words or phrases in the title and or abstract:

- anthropogenic
- climate action
- climate anxiety
- climate change
- climate crisis
- climate resilient
- COP (conference of the parties) meetings
- environmental sustainability
- global warming
- greenhouse effect
- greenhouse gases
- nationally determined contributions (NDCs)
- nature crisis or planetary crisis
- planetary health.

Include articles that:

- refer to greening of health systems, or any indication of progressive change in the climate over time (even if this is not the main focus of the article)
- refer to carbon footprinting
- refer to making changes in health care to make it more environmentally sustainable
- refer to disinvesting from fossil fuels or taking other action to reduce the use of fossil fuels
- include the word biodiversity after you have checked that the article is covering threats to biodiversity or ways of enhancing it. (Biodiversity seems to be a term that has become widely used in the context of the planetary crisis, and almost all articles that include the word are likely to be included)
- are about animal and plant (not just human) health & climate change.

#### Exclusion criteria

Do NOT include articles:

- on the consequences of climate change - like, for example, heatwaves, floods, wildfires, storms, other weather extremes, or changing patterns in infectious disease - that do NOT mention changing climate. These phenomena happened before climate change began but are being made more common by climate change
- on air pollution unless they include specific references to climate change (using the full range of terms)
- that refer simply to seasonal change in climate
- that refer to changes in disease patterns because of geographical changes in climate—for example, asthma in people who move geographically from one climate to another
- that refer to changes in climate of opinion, political climate, or any reference to climate that is not associated with weather.

## REFERENCES

1. Costello A, Abbas M, Allen A, et al. Lancet and University College London Institute for Global Health Commission. Managing the health effects of climate change. Lancet 2009; 373: 1693–1733. doi:10.1016/S0140-6736(09)60935-1

2. Wang H, Horton R. Tackling climate change: the greatest opportunity for global health. Lancet 2015; 386:1798-9. doi:10.1016/S0140-6736(15)60931-X

3. Romanello M, Di Napoli C, Drummond P, et al. The 2022 report of the Lancet Countdown on health and climate change: health at the mercy of fossil fuels. Lancet 2022; 400: 1619–54. doi:10.1016/S0140-6736(22)01540-9

4. Health Care Without Harm and Arup. Health Care’s Climate Footprint: How the health sector contributes to the global climate crisis and opportunities for action. Health Care Without Harm and Arup. 2019. https://noharm-global.org/sites/default/files/documents-files/5961/HealthCaresClimateFootprint_092319.pdf (Accessed 18 October 2023).

5. NHS England. Delivering a ‘Net Zero’ National Health Service. London: NHS England, 2019. https://www.england.nhs.uk/greenernhs/wp-content/uploads/sites/51/2022/07/B1728-delivering-a-net-zero-nhs-july-2022.pdf (Accessed 18 October 2023).

6. Krajick K. James Hansen’s Climate Warning, 30 Years Later. Columbia Climate School. 2018; 26 June. https://news.climate.columbia.edu/2018/06/26/james-hansens-climate-warning-30-years-later/ (Accessed 18 October 2023).

7. Berrang-Ford L, Sietsma AJ, Callaghan M, et al. Systematic mapping of global research on climate and health: a machine learning review. Lancet Planet Health 2021;5(8):e514–e525. doi:10.1016/S2542-5196(21)00179-0

8. Verner G, Schütte S, Knop J, et al. Health in climate change research from 1990 to 2014: positive trend, but still underperforming. Global Health Action 2016; 9:1. doi:10.3402/gha.v9.30723

9. Harper SL, Cunsolo A, Babujee A, et al. Climate change and health in North America: literature review protocol. Syst Rev 2021;10(1):3. doi:10.1186/s13643-020-01543-y

10. Bartlett VL, Gupta R, Wallach J, et al. Published Research on the Human Health Implications of Climate Change from 2012-2019: a Cross-Sectional Study. MedRxiv 2022;10.01.22280596. doi:10.1101/2022.10.01.22280596

11. Atwoli L, Baqui AH, Benfield T, et al. Call for emergency action to limit global temperature increases, restore biodiversity, and protect health. BMJ 2021;374:n1734. doi:10.1136/bmj.n1734.

12. Atwoli L, Erhabor G E, Gbakima A A, et al. COP27 climate change conference: urgent action needed for Africa and the world BMJ 2022; 379 :o2459. doi:10.1136/bmj.o2459

13. Einecker R, Kirby A. Climate Change: A Bibliometric Study of Adaptation, Mitigation and Resilience. Sustainability 2020; 12(17):6935. doi:10.3390/su12176935

14. Fu HZ, Waltman L. A large-scale bibliometric analysis of global climate change research between 2001 and 2018. Climatic Change 2022;170, 36. doi:10.1007/s10584-022-03324-z

15. Sangam SL, Savitha KS. Climate change and global warming: A scientometric study, COLLNET Journal of Scientometrics and Information Management 2019; 13(1) 199–212. doi:10.1080/09737766.2019.1598001

16. Sweileh, WM. Bibliometric analysis of peer-reviewed literature on climate change and human health with an emphasis on infectious diseases. Global Health 2020;16, 44. doi:10.1186/s12992-020-00576-1

17. Sweileh, WM. Bibliometric analysis of peer-reviewed literature on food security in the context of climate change from 1980 to 2019. Agric & Food Secur 2020; 9,11. doi:10.1186/s40066-020-00266-6

18. Li F, Zhou H, Huang D-S, Guan P. Global Research Output and Theme Trends on Climate Change and Infectious Diseases: A Retrospective Bibliometric and Co-Word Biclustering Investigation of Papers Indexed in PubMed (1999–2018). International Journal of Environmental Research and Public Health 2020; 17(14):5228. doi:10.3390/ijerph17145228

19. Neergaard KV. Dangers of climate change. [in German] Schweiz Med Wochenschr 1947;77(44):1160.

20. Hollander JL. [The effect of climate changes upon the evolution of rheumatoid arthritis]. [in Spanish] Rev Med Chil 1964;92:180–3.

21. Cuenot A. [Psychism and change of climate]. Presse Med 1961;69:35–8.

22. Ebi KL, Semenza JC, Rocklöv J. Current medical research funding and frameworks are insufficient to address the health risks of global environmental change. Environ Health 2016;15, 108. doi:10.1186/s12940-016-0183-3

23. Hai Z, Perlman RL. Extreme weather events and the politics of climate change attribution. Sci Adv 2022;8(36):eabo2190. doi:10.1126/sciadv.abo2190

